# Gluteus Maximus Shape Reveals Sex-specific Associations between Morphology and Metabolic Dysfunction

**DOI:** 10.64898/2026.04.01.26349946

**Authors:** Marjola Thanaj, Brandon Whitcher, Hamzah Raza, Camilo Bradford-Bell, Marili Niglas, Jimmy D Bell, Dimitri Amiras, E Louise Thomas

## Abstract

**Background:** The gluteus maximus (GM) is a major hip extensor essential for mobility and metabolic health. Most MRI studies rely on global measures, such as muscle volume or fat fraction, which can overlook spatially localised remodelling. Here, we integrate conventional volumetric and fat fraction metrics with 3D mesh-based shape phenotypes to provide a spatially resolved characterisation of GM morphology in relation to anthropometric, lifestyle, and cardiometabolic factors, with a focus on type 2 diabetes (T2D) and sex-specific effects.

**Methods:** We analysed T1 Dixon MRI from UK Biobank participants to quantify GM muscle volume, fat fraction, and regional surface morphology using 3D meshes. Statistical parametric mapping was used to assess regional associations with anthropometric, lifestyle, and clinical variables Bi-directional causal mediation analyses were performed using GM volumetric and principal components (PCs) of shape variation. PCs were also tested for associations with prevalent and incident disease. Longitudinal changes in GM composition were evaluated in participants with repeated imaging evaluations.

**Results:** GM muscle volume and fat fraction were strongly associated with age, adiposity, and physical activity. Shape analysis revealed spatially localised remodelling patterns not captured by global measures, with region-specific surface shrinkage linked to age, BMI, alcohol intake, grip strength, physical activity, frailty, osteoporosis, and cardiometabolic disease. T2D showed marked sex-differences, with regional shrinkage in men and relative expansion in women. PCA reduced high-dimensional shape variation into interpretable components. Mediation analyses indicated that T2D-related differences in GM morphology partly mediated increases in fat fraction, suggesting that disease effects manifest through spatially patterned shape changes rather than overall muscle size. PCs capturing variations in the central-upper posterior and anterior GM, differentiated between T2D cases from controls, and were associated with incident T2D risk (Men: PC6 HR per SD: 0.81 [0.70–0.95], false discovery rate (FDR)-adjusted p = 0.038, in left GM; 0.76 [0.65–0.88], p = 0.002, in right GM; women; PC5 HR= 1.32, [1.08–1.61], p = 0.032, in right GM).

**Conclusions:** Integrated 3D quantification of GM composition and morphology provides spatially resolved biomarkers that go beyond muscle volume and fat fraction. By capturing region-specific GM remodelling, linked to anthropometric, lifestyle and cardiometabolic factors, this approach offers a more nuanced characterisation of muscle–fat phenotypes and enhances mechanistic insight and risk stratification in population-based imaging studies.

## Introduction

Skeletal muscle composition is a key determinant of mobility and metabolic health, with age- and disease-related fat infiltration (myosteatosis) increasingly recognised as an imaging marker of impaired muscle quality^1^. In the hip extensors, the gluteus maximus (GM) is one of the body’s largest muscles and is central to pelvic stability, posture, and high-intensity tasks such as climbing, sprinting, and lifting^2^. A growing literature using CT and MRI has linked muscle fatty degeneration to poorer function and clinical outcomes, highlighting that compositional change can be more informative than muscle size alone^3^. In the GM specifically, higher fat content has been associated with reduced mobility, greater fall risk, and poorer performance^4^, while population studies have shown that low physical activity is related to GM intramuscular fat^5^.

These findings align with broader evidence that accumulation of ectopic fat is closely connected to metabolic dysfunction^6^. Individuals with type 2 diabetes (T2D) show distinct adipose tissue distribution patterns^7^, and sex-related differences in body composition and insulin resistance may contribute to heterogeneity in metabolic risk^8^. However, existing work on GM measurements has typically been conducted in relatively small or disease-specific cohorts and has largely focused on global summaries (e.g., mean fat fraction or total volume), leaving important gaps. First, the relationships between GM muscle volume, fat fraction, a wide range of lifestyle and anthropometric factors, and both prevalent and future disease risk, have not been characterised at scale within a single harmonised imaging framework. Second, global measures cannot pinpoint where clinically relevant remodelling occurs within the GM, which limits biological interpretability and potentially obscures spatially specific signatures of metabolic dysfunction.

Three-dimensional (3D) mesh-derived phenotypes capture morphological and regional variation through statistical parametric maps (SPMs). Statistical shape analysis (SSA) further summarises spatially correlated data into principal components that characterise population-level shape variation independent of volume. MRI studies traditionally focus on measuring volumes or fat infiltration, whereas statistical shape analysis characterises tissue morphology and localises shape variation via statistical parametric maps (SPMs), independent of volume. While this is widely used to model bones^9^, abdominal organs^10,11^, the brain^12^, the heart^13^ and the aorta^14^, its application to skeletal muscle, especially the GM, remains limited.

In this study, we analyse T1 Dixon MRIs from the UK Biobank to quantify GM muscle volume, intermuscular adipose tissue (IMAT), muscle fat fraction, and regional surface morphology. Our aims were to: (i) characterise cross-sectional associations between GM muscle-fat phenotypes and anthropometric and lifestyle factors; (ii) assess associations between GM phenotypes and prevalent and incident disease, focusing on T2D and sex-specific effects; (iii) quantify longitudinal changes using repeat imaging; (iv) investigate interrelationships between GM morphology, T2D, and fat fraction; and (v) determine whether 3D mesh-derived phenotypes improve T2D diagnosis and prediction of future disease risk.

## Methods

### Data

Full details regarding the UK Biobank abdominal MRI acquisition protocol have previously been reported^15^. Briefly, the data included here focused on neck-to-knee Dixon MRI acquisitions, comprising six overlapping series. Data were obtained under UK Biobank applications 44584 and 23889. The UK Biobank has approval from the North West Multi-Centre Research Ethics Committee (REC reference: 11/NW/0382). All methods were performed in accordance with relevant guidelines, with informed consent obtained from all participants. Further information is available at www.ukbiobank.ac.uk.

### Image and Mesh-Derived Analysis of GM

#### Image Analysis

All Dixon MRI data were preprocessed using previously described methods^16^. We utilised a three-dimensional (3D) U-net model architecture in the MONAI (Medical Open Network for AI) framework for segmentation^17^. Manual annotation of the GM muscles was performed on 158 datasets. Data augmentation was performed, and a total of 384 epochs were run; the final model was selected using the Dice score on 10% out-of-sample datasets. Left/right separation of the GM was performed in post-processing. The pipeline was applied to over 58,000 neck-to-knee T1-weighted Dixon MRI scans, with segmentation quality validated by experienced analysts at multiple stages.

IMAT was identified from Dixon fat images thresholded at a 0.5 fat fraction cut-off, with remaining tissue classified as muscle (Supplementary Figure S1). The fat/muscle ratio was calculated as: GM IMAT volume / (GM IMAT volume + GM muscle volume). Image-derived phenotypes (IDPs) included GM muscle volume, IMAT volume, and IMAT/muscle ratio.

#### Image Registration and Mesh Construction

Population-based templates^18,19^ were constructed separately for left/right GM in 600 participants (300 men, 300 women). Template characteristics and exclusions are provided in Supplementary Table S1, and in Data SD1. 3D surface meshes were constructed from template and participant segmentations using the marching cubes algorithm. Rigid registration^18,19^ was used to remove positional and orientation differences between all participant-specific surfaces and the template surfaces, followed by affine and non-rigid registration of the template to each participant’s segmentation. The template mesh was propagated to each participant using the resulting deformation fields, yielding meshes parameterised with approximately 16,000 vertices (men) and 13,000 vertices (women), ensuring anatomical consistency across all participants while preserving size and shape information.

Surface-to-surface (S2S) distances, the 3D mesh-derived phenotype were measured as the signed distance between each template vertex and the corresponding participant vertex. Positive distances represent outward expansion while negative distances indicate inward shrinkage relative to the template^19^ (Supplementary Figure S2).

### Quality Control

The study included MRI data from 55,864 participants at initial imaging visit and 2,912 participants with a follow-up scan approximately 2.5 years later^20^. We excluded 7,826 participants with missing anthropometric, biological, or lifestyle variables, and four with zero GM volume. Outliers in the 3D mesh-derived phenotype were inspected visually; no further exclusions were required. The final baseline sample comprised 48,034 participants. For the longitudinal sample, 198 participants were excluded due to missing data, yielding 2,720 participants (Supplementary Figure S3).

### Statistical Analysis

#### Model Selection

We fitted least absolute shrinkage and selection operator (LASSO) regression models, implemented in the R package *glmnet*, via a stability selection procedure using the R package *stabs*^21^ selecting models across 85 demographic, biomarker, and disease data sets (Supplementary Data SD2). Stability selection was performed using the “stabsel” function with the fitting function set to “glmnet.lasso”. Stability selection used a probability cutoff of 0.95 and per-family error rate of one, wth unimodal assumption. The stability parameters were estimated using the “stabsel_parameters” function, ensuring a robust selection of predictors^21^. Model-based boosting was implemented using the package *mboost*^22^.

#### Linear Regression Analysis

Selected covariates were entered into linear regression models to assess associations with GM IDPs. Longitudinal analyses in 2,720 participants used linear mixed-effects models fitted with *lme4* package^23^, included participant IDs as a random effect, T2D as a fixed effect, including data from both time points^24^. Models further included stability-selected covariates, assessment centre, imaging visit, T2D, and their interaction. P-values were computed using *lmerTest* package^25^ with FDR correction (threshold <0.05). Effect sizes are reported as standardised beta coefficients (β). All continuous variables were standardised prior to being included in the models.

#### Mass Univariate Regression Analysis

Associations between S2S distances and anthropometric variables were estimated using a mass univariate regression (MUR) framework. We applied threshold-free cluster enhancement (TFCE)^26^ and permutation testing, with 1,000 permutations and FDR correction, as previously described^18^ (see Supplementary material and Figure S4).

Sex-stratified linear regression was fitted at each vertex, adjusting for stability-selected covariates. Interaction terms between age and T2D were included to assess accelerated change. All continuous variables, including S2S distances, were standardised prior to analysis.

Longitudinal S2S analyses in 2,720 participants applied the same MURr mixed-effects framework with TFCE with 1,000 permutation testing and FDR correction, and a threshold of < 0.05 determined significance. All continuous variables used as fixed effects were standardised before being included in the analysis. Wilcoxon rank-sum tests were used for paired comparisons between visits.

#### Dimensionality Reduction of GM Shape

To reduce the dimensionality of the 3D mesh-derived phenotypes, we computed robust sparse principal component analysis (SPCA) using the R package *sparsepca*^27^ and extracted principal component (PC) scores to characterise variations in GM S2S distances for each gender and each GM. Modes of variation (−3 SD, mean, +3 SD) were mapped onto representative templates for visualisation.

#### Causal Mediation Analysis

Bidirectional causal mediation analyses were performed separately by sex and GM side to decompose T2D-related effects on GM fat fraction into direct and indirect components. Analyses considered GM muscle volume and the first ten principal component (PC) scores derived from PCA of the S2S distances (capturing the major modes of GM shape variation). Specifically, we evaluated: (i) T2D as exposure, GM muscle volume as mediator, GM fat fraction as outcome; (ii) T2D as exposure, each PC score as mediator, GM fat fraction as outcome; (iii) GM muscle volume as exposure, T2D as mediator, GM fat fraction as outcome and (iv) each PC score as exposure, T2D as mediator, GM fat fraction as outcome. All models were adjusted for stability-selected covariates, with continuous variables standardised prior to analysis. The average causal mediation effect (ACME), average direct effect (ADE), and total effect were estimated using nonparametric bootstrap resampling (5,000 simulations) and p-values□<□ 0.05 were considered statistical significant. Mediation analyses used the R package *mediation*^28^.

#### Diagnostic Modelling

To evaluate whether GM PCs improved T2D diagnosis beyond conventional IDPs, we identified prevalent T2D cases and matched controls (1:1) by age (±2 years) and BMI (±1.5 kg/m²) using *ccoptimalmatch*^29^ stratified by sex. Sex-specific logistic regression models included bilateral GM volume, fat fraction, and PC scores, alongside non-disease stability-selected covariates. The dataset was split by assessment centre into training (Cheadle, Bristol, Newcastle) and test (Reading) subsets. Models were trained using repeated 10-fold cross-validation (five repeats) with the R package *caret*^30^.

Two nested models were compared: a baseline model with GM volume and fat fraction, and an extended model additionally incorporating PC2–PC10 of S2S distances, excluding PC1 explains the size of GM to avoid redundancy with muscle volume. Performance was assessed using AUC, accuracy, sensitivity and specificity. Odds ratios (ORs) with 95% confidence intervals were derived from the final fitted models, and multiple testing was controlled using FDR correction.

#### Survival Analysis

Cox proportional hazards models evaluated incident T2D risk from the imaging visit, fitted separately by GM side and sex, adjusted for stability-selected covariates measured at the imaging visit. To minimise overfitting given limited events, penalised LASSO-Cox regression was applied. dataset was split by assessment centre into a training set (Cheadle, Bristol, Newcastle) and an independent test set (Reading). LASSO selection was conducted in the training data using 10-fold cross-validation to identify the optimal penalty parameter. Variables retained by LASSO were entered into standard Cox models to estimate hazard ratios (HRs). Model discrimination was assessed using concordance indices (c-index) in the held-out test set.

Disease outcomes were defined using linked hospital records, primary care data, self-reports, and death registries (see Supplemetary material). Follow-up was calculated from the imaging visit and censored at first event, death, or 30 November 2022 (median 4 years). Participants with prior T2D were excluded. Hazard ratios (HRs) with 95% confidence intervals were reported, with FDR-adjusted p < 0.05 indicating statistical significance. Survival analyses used the R package *survival*^31^, and all continuous variables were standardised prior to analysis.

## Results

### Segmentation Performance

The deep learning model achieved a mean Dice similarity coefficient of 0.94 on a held-out 10% test set (15 participants). Visual inspection confirmed successful segmentation.

### Study Population Characteristics

The cohort included 23,364 men and 24,670 women. Among men, 96.6% were White, ages ranged from 44 to 85 years, and mean BMI was 27 ± 3.9 kg/m². In women, 96.8% were White, ages ranged from 45 to 84 years, and mean BMI was 26.1 ± 4.8 kg/m² (Supplementary Table S2). We identified 2,628 participants with T2D (1,699 men; 896 women), 1,140 with frailty (472 men; 668 women), 3,917 with osteoporosis (1,034 men; 2,883 women), and 17,615 with CVD, of whom 58% were men.

### Associations with Anthropometric Traits and Disease

We examined associations between GM muscle volume, IMAT volume, and fat fraction with covariates selected by the stability selection model, including age, ethnicity, BMI, waist-to-hip ratio (WHR), alcohol intake, dominant handgrip strength (HGS), vigorous metabolic equivalent task (MET), serum creatinine, cystatin C, urinary creatinine, IGF-1, white blood cell count (WBC), and clinical conditions including T2D, frailty, osteoporosis, and CVD (Supplementary Figure S5). We further adjusted for time since the initial assessment visit to account for the time gap between the initial assessment visit, where the biomarkers were taken, and the imaging visit (Supplementary Figure S6 and Table S3). Effect sizes are reported as standardised beta coefficients (β) with FDR-adjusted p-values.

GM muscle volume was negatively associated with age, cystatin C, WBC, frailty, CVD, and osteoporosis in both sexes. In men, T2D was additionally associated with lower muscle volume (left: β = –0.14, p = 1.9×10□ ¹³; right: β = –0.11, p = 5.3×10□□). By contrast, dominant HGS and vigorous physical activity were positively associated with GM muscle volume. IMAT volume and fat fraction were positively associated with BMI, alcohol intake, IGF-1, T2D, frailty, and osteoporosis, while HGS and physical activity were inversely associated with fat fraction. Age × T2D interactions showed sex-specific patterns: in women, older age was associated with lower GM muscle and IMAT volume (left muscle: β = –0.10, p = 0.028; right muscle: β = –0.10, p = 0.015; left IMAT: β = –0.09, p = 1.8×10□□ ; right IMAT: β = –0.12, p = 5.5×10□□), whereas in men, age was positively associated with GM fat fraction (left: β = 0.15, p = 0.045; right: β = 0.14, p = 0.040).

MUR analysis of S2S distances generated SPMs adjusted for all relevant covariates (Table 1-2, Supplementary Figure S7). Age was predominantly negatively associated with S2S distances in both sexes (men: 71–75% of GM surface; women: 71% bilaterally), indicating inward shape variation. BMI showed positive associations across nearly the entire GM surface (men: 98.7% left, 99.6% right; women: 97.7% left, 98.6% right). WHR patterns differed by sex: men showed mainly negative associations, while women exhibited a mixed pattern, with positive associations covering 54% (left) and 58% (right) of the lateral-to-inferior GM and negative associations in smaller superior and iliosacral areas. Serum creatinine, IGF-1, urinary creatinine, vigorous activity, and HGS were positively associated with S2S distances across >75% of GM. Cystatin C and WBC were negatively associated, particularly in men. Alcohol intake showed predominantly positive associations.

T2D was associated with inward S2S displacements in men and outward displacements in women, indicating opposite deformation patterns across the GM surface. To facilitate interpretation, unstandardised regression coefficients translating shape variation into mm are provided in Supplementary Table S4. Based on the median SD of S2S distances in men (4.74 mm bilaterally), T2D corresponded to median inward shape variation in the superior and inferior posterior areas and lateral-to-inferior anterior areas with a median of –0.38 mm (left) and –0.33 mm (right) in men, and outward variation in the lateral-to-inferior (posterior and anterior) GM with a median of 0.42 mm (left) and 0.40 mm (right) in women. Frailty showed widespread inward deformation in men (–0.47 mm left, –0.71 mm right) and more heterogeneous effects in women, with inward deformations across roughly half the GM (–0.37 mm left, –0.36 mm right) and localised outward deformations. Osteoporosis was consistently associated with inward deformation in both sexes (men: – 0.66 mm bilaterally; women: –0.37 mm left, –0.45 mm right). CVD was associated with predominantly inward remodelling across large areas of both GM surfaces, with larger effects in men (–0.24 mm bilaterally) than women (–0.12 mm left, –0.13 mm right) (Supplementary Videos S1).

Age × T2D interaction analysis showed that age-related inward deformation was more pronounced in participants with T2D in both sexes. In men, age was associated with stronger inward GM deformation particularly in the anterior right GM, with an additional median S2S distances of –0.14 vs –0.08 (right) for T2D vs non-T2D participants while in women, median S2S distances were –0.14 vs –0.07 in both GM, affecting mainly central posterior and lateral anterior regions (Table 2, Figure 1-2). Interactions between T2D and GM fat fraction revealed stronger outward shape variation in those with higher fat fraction in men (left: 0.10 vs 0.05; right: 0.11 vs 0.06), affecting 33% of the left and 55% of the right GM, with the largest expansion in the central-posterior right GM. Similar patterns were observed in women (left: 0.09 vs 0.05; right: 0.12 vs 0.05), affecting 54% of the left and 21% of the right GM, with regional predominance in the central left and central-anterior right GM border with the gluteus medius and minimus (Supplementary Figure S8).

**Figure 1.**
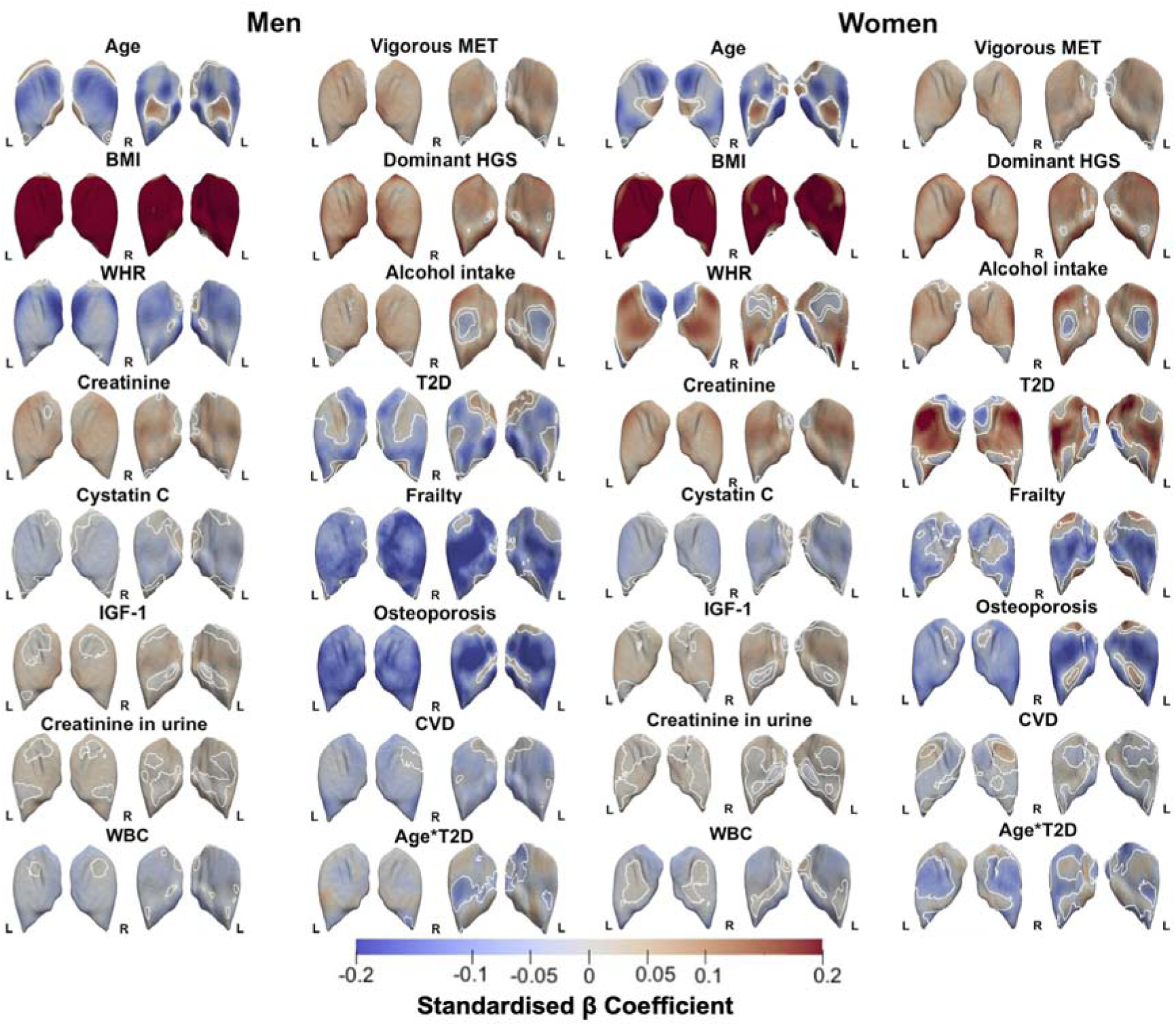
Three-dimensional sex-specific SPMs of GM morphology, projections are for both left (L) and right (R) GM in both posterior (left plots) and anterior (right plots) views. The SPMs show the local strength of association for each covariate in the model with S2S distances in men (N=23,364) and women (N=24,670). White contour lines indicate the boundary between statistically significant regions (p < 0.05) after correction for multiple testing, with bright red showing an outward deformation (fatty hypertrophy) and bright blue showing inward deformation (atrophy). The standardised regression coefficients (r) are shown with units in standard deviations for each covariate. Abbreviations: SPM: statistical parametric map; GM: gluteus maximus; S2S: surface-to-surface; BMI: body mass index; WHR: waist-to-hip ratio, IGF-1: insulin-like growth factor 1; WBC: white blood cell count; MET: metabolic equivalent task; HGS: hand grip strength; T2D: type-2 diabetes; CVD: cardiovascular disease.

**Figure 2.**
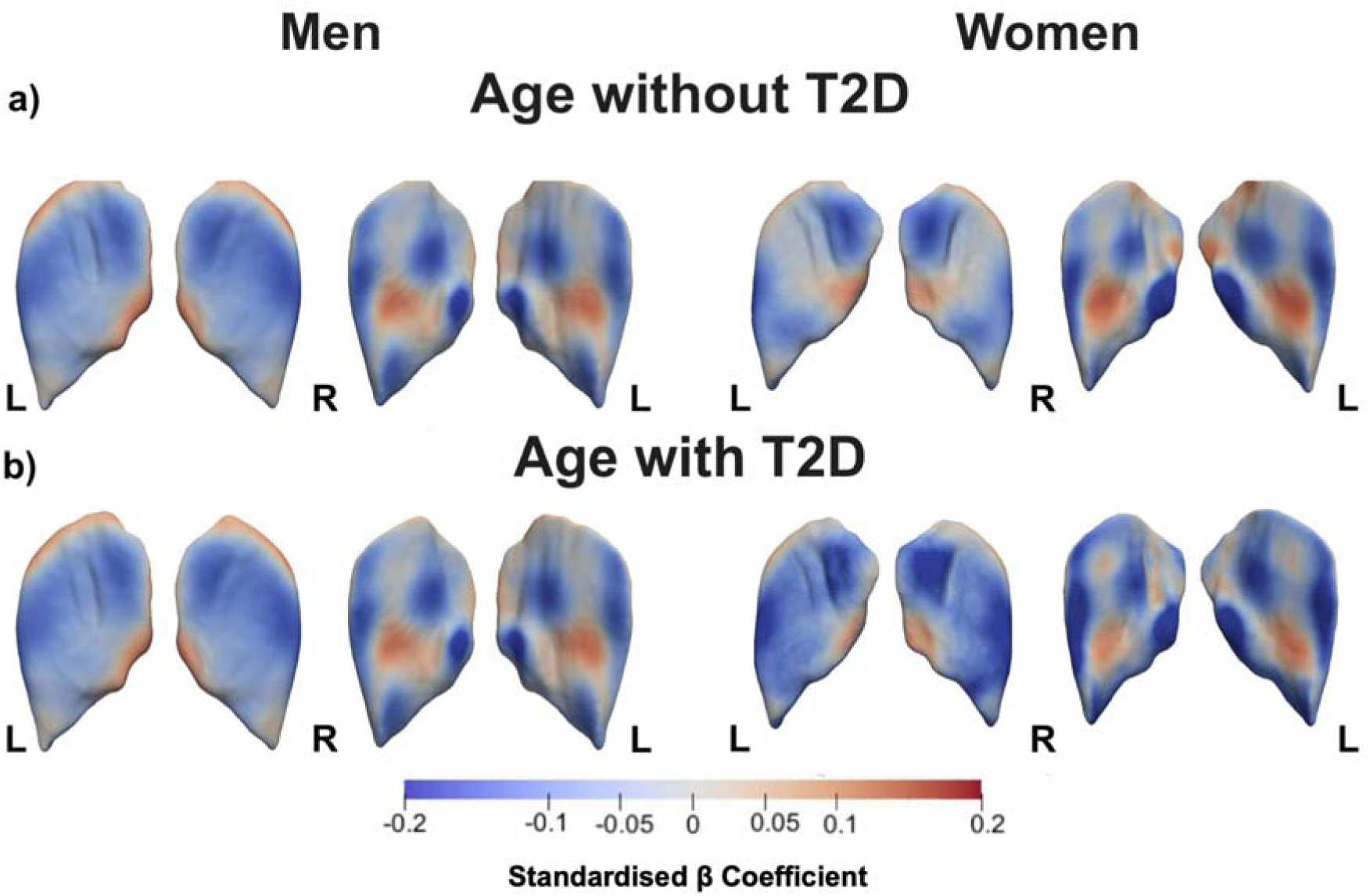
Three-dimensional sex-specific SPMs of GM morphology, projections are for both left (L) and right (R) GM in both posterior (left plots) and anterior (right plots) views. The SPMs show the local rate of change as a function of age for S2S distances in participants **a)** without T2D and **b)** with T2D, referred to the sums of the regression coefficients for Age and Age × T2D, in men (N=23,364) and women (N=24,670). Positive associations are in red and negative associations are in blue. Regression coefficients (r) are shown with units in standard deviations for each covariate. Abbreviations: SPM: statistical parametric map; GM: gluteus maximus; S2S: surface-to-surface; T2D: type-2 diabetes.

### Longitudinal Changes in GM

Among 2,720 participants with repeat imaging after 2.25 ± 0.12 years (1,362 men; 1,358 women), sex-stratified linear mixed-effects models adjusted for age, assessment centre, ethnicity, BMI, WHR, alcohol intake, HGS, vigorous MET, and T2D × visit interaction revealed consistent longitudinal changes across sexes (Supplementary Figures S9–S10). Independent of T2D status, the second imaging visit was associated with declines in GM muscle volume and increases in IMAT volume and fat fraction. Women showed decreases in muscle volume and increases in IMAT and fat fraction while men exhibited greater muscle loss and larger increases in IMAT and fat fraction (Supplementary Table S5).

Longitudinal changes using mass univariate linear mixed-effect models in GM S2S distances showed no significant impact of T2D on overall shape trajectories. However, both sexes exhibited notable regional shape changes at follow-up (Figure 3). In men, median S2S distances decreased by 0.49 mm (left, 41.5% significance area) and 0.51 mm (right, 32.6%) in inferior posterior/anterior areas and in the sciatic notch, with increases of 0.30 mm (left, 42.7%) and 0.37 mm (right, 28.4%) in central posterior and inferior/lateral anterior regions (Supplementary Tables S6–S7). Similar median longitudinal differences were observed in women (Supplementary Figure S11).

**Figure 3.**
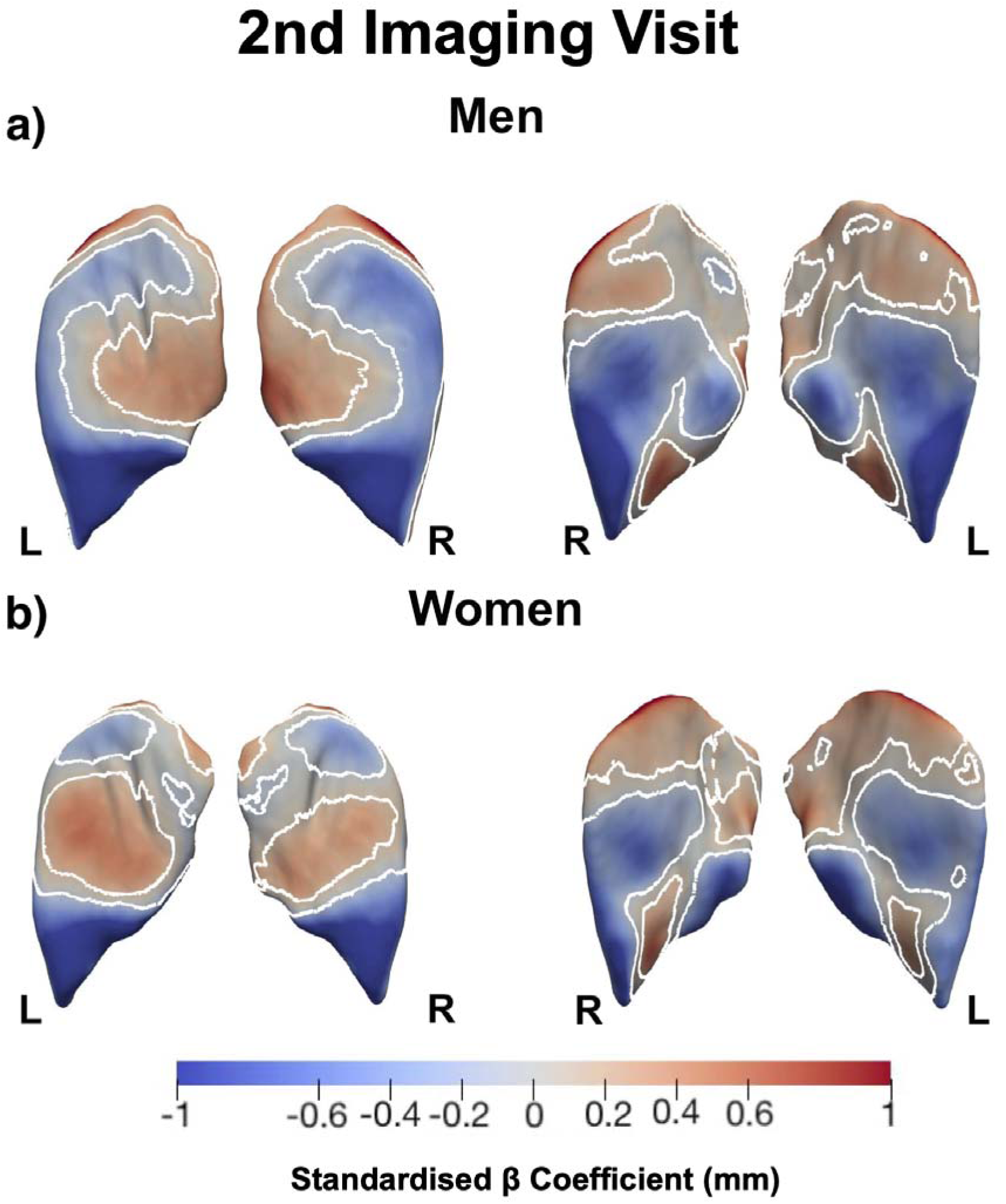
Three-dimensional sex-specific SPMs of GM morphology, projections are for both left (L) and right (R) GM in both posterior (left plots) and anterior (right plots) views. The SPMs show the local strength of association of S2S distances and the imaging visit for **a)** men (N=1,362) and **b)** women (N=1,358). White contour lines indicate the boundary between statistically significant regions (p < 0.05) after correction for multiple testing, with positive associations in bright red and negative associations in bright blue. The standardised regression coefficients (β̂) are shown with units in standard deviations for each covariate. Abbreviations: SPM: statistical parametric map; GM: gluteus maximus; S2S: surface-to-surface.

### Mediation effects in the association of GM morphology, T2D and GM fat fraction

SPCA captured over 70% of S2S variation across the first ten PCs in both sexes (Supplementary Figure S12 and Video S2).

In men, GM muscle volume showed significant indirect effects on fat fraction when mediated by T2D (left: ACME = −0.003, p < 1×10⁻¹⁶; right: ACME = −0.003, p = 0.001), with strong direct effects remaining (left: ADE = −0.150, p < 1×10⁻¹⁶; right: ADE = −0.100, p < 1×10⁻¹⁶). Conversely, when T2D was the exposure and GM muscle volume the mediator, significant indirect effects were also observed (left: ACME = 0.022, p < 1×10⁻¹⁶; right: ACME = 0.012, p < 1×10⁻¹⁶). In women, no significant indirect effects on muscle volume were observed (Supplementary Figures S13–S14).

PC-based mediation revealed sex-specific effects. In men, T2D was negatively associated with PC1 (reflecting smaller GM size), while PC1 was positively associated with GM fat fraction, producing a significant negative indirect effect that slightly attenuated the total T2D–fat fraction association, with the direct effect remaining dominant (Supplementary Figure S15). PC3, representing lateral expansion and shrinkage in supero-inferior regions, significantly mediated the T2D–fat fraction association in the right GM (ACME = 0.003, p = 0.005) but not the left GM. In the left GM, PC4 capturing central/superior/inferior expansion and shrinkage in the sciatic notch, and PC7 reflecting femoral region shrinkage and central/superior expansion, showed significant mediation effects (PC4: p = 0.016; PC7: p = 0.006; Supplementary Figures S16–S18).

In women, PC1 showed no significant indirect effects through T2D in either direction (p > 0.05, Figure 4a–b). However, when T2D was the exposure and PC1 the mediator, small but significant indirect effects were observed bilaterally (left: ACME = 0.003, p = 0.02; right: ACME = 0.004, p = 0.006), indicating that part of the T2D-related increase in fat fraction operates through GM size variation (Figure 4c–e). PC2, explaining superior/inferior and ilioscral expansion and central/femoral region shrinkage, significantly mediated the T2D–left GM fat fraction association in both directions (as exposure: ACME = −0.001, p < 0.05; as mediator: ACME = 0.005, p = 8×10□□), with no significant effects observed in the right GM (Supplementary Figure S19).

**Figure 4.**
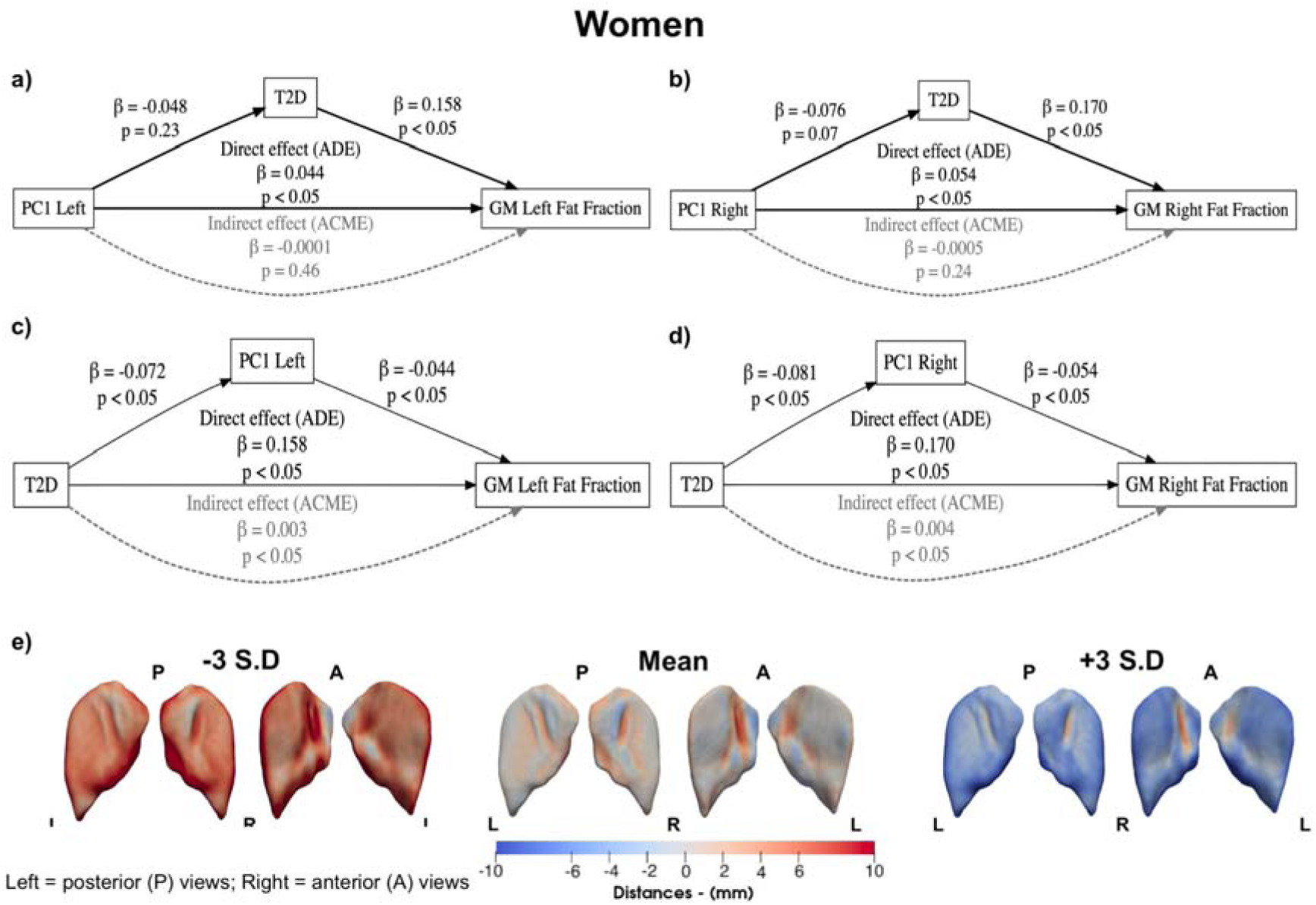
Causal mediation analysis on the relationship between PC1 explaining GM size (from larger to smaller), T2D and GM fat fraction in women (N=24,670). Upper panels **a)** and **b)** show exposure PC1 of GM left and right with mediator T2D. Lower panels **c)** and **d)** show exposure T2D with mediator PC1 of GM left and right. **e)** The mean shape and the shape at the ±3 SD are displayed for PC1, showing the S2S distance variation in mm. The GM shape variations are shown in the anterior and posterior views of each GM. Direct effects are indicated by solid black arrows, whereas mediation effects are indicated by dashed grey arrows. Labels indicate effect size and p-values. Abbreviations: PC: principal component; GM: gluteus maximus; S2S: surface-to-surface; T2D: type-2 diabetes.

### Diagnostic and Survival Analysis

Two models were compared in T2D case-control cohorts (men N=2,058; women N=1,318) using 81%/78% training and 19%/21% test splits: a volume model including GM volume, fat fraction, and non-disease stability-selected covariates, and an extended volume + PCs model additionally incorporating PC2–PC10 of GM S2S distances (Supplementary Figures S20–S21).

In men, the volume model achieved AUC=0.77 for both GM, with a modest improvement on adding PCs (AUC=0.78). T2D was associated with higher GM fat fraction (left OR = 1.56 [1.36–1.81], p = 1.4×10□□; right OR = 1.61 [1.40–1.86], p = 1×10□□) and PC6 reflecting lateral and inferior GM variations (left OR = 0.85 [0.77–0.95], p = 0.015; right OR = 1.21 [1.08–1.35], p = 0.002). In women, the volume model achieved AUC = 0.75, increasing to 0.76 with the addition of PCs for both GM. T2D was associated with lower PC5 and PC6 in the left GM, capturing central shrinkage with sciatic notch expansion.

Cox proportional hazards models were used to assess the risk of incident T2D after the imaging visit, adjusting selected covariates measured at the imaging visit, GM volume, fat fraction, and PC2-PC10 (Supplementary Figure S22). Data were split into 84%/83.5% training and 16%/16.5% testing for men and women, respectively. Models demonstrated good discrimination (C-index: men 0.82 bilaterally; women 0.74 left, 0.76 right). In men, higher PC6, reflecting reduced variations in the central-upper GM, was consistently associated with lower T2D risk (left HR = 0.81 [0.70–0.95], FDR-adjusted p = 0.038; right HR = 0.76, 95% CI 0.65–0.88, p-value = 0.002). In women, PC5 of the right GM, encompassing the central posterior and upper anterior GM, was significantly associated with increased T2D risk (HR = 1.32 [1.08–1.61], p = 0.032) (Figure 5).

**Figure 5.**
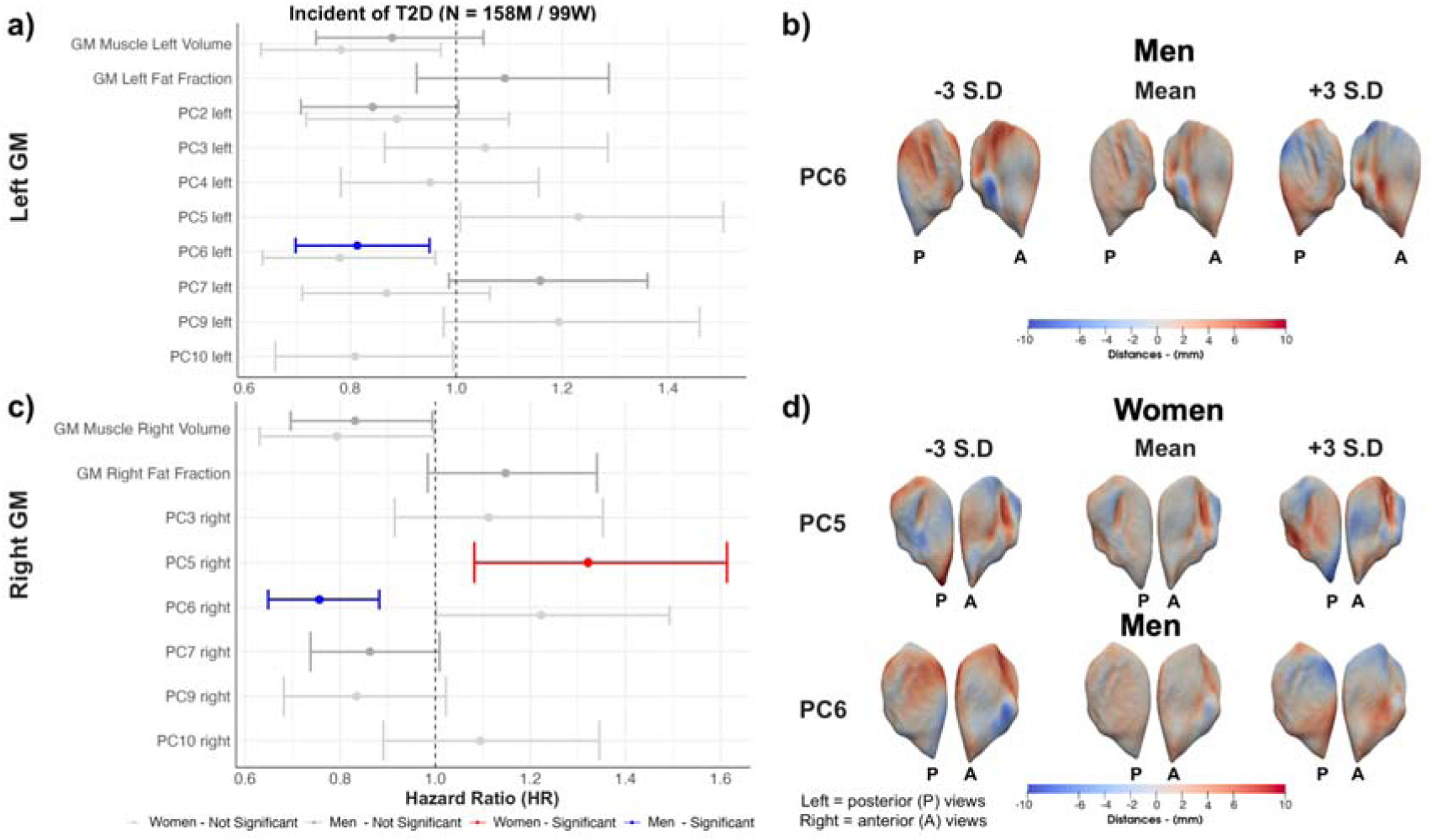
Hazard ratios and 95% CIs for T2D outcome for **a)** the men (N =158) and **c)** women (N = 99) participants in both left and right GM, adjusted for variables selected from the LASSO selection, including anthropometric, lifestyle factors and the first ten PC scores for GM S2S distances. Statistically significant associations (FDR-adjusted p < 0.05) are shown in red. The PCs are visualised for **b)** men and **d)** women, representing the significant PCs of shape variation for the left and right GM. The mean GM shape and the shapes at ±3 SD are displayed for each PC, showing the S2S distance variation in mm. The GM shape variations are shown in the anterior and posterior views of each GM. Abbreviations: LASSO: least absolute shrinkage and selection operator; T2D: type-2 diabetes; GM: gluteus maximus; S2S: surface-to-surface; PC: principal component; FDR: false discovery rate.

## Discussion

This study integrates conventional MRI-derived GM IDPs with mesh-based morphometry to characterise global composition and regional remodelling in the UK Biobank imaging cohort. Combining cross-sectional associations, longitudinal analyses, mediation modelling, and prediction and survival analyses, we show that GM fat fraction and localised shape patterns act as complementary markers of metabolic and functional health.

IMAT and fat fraction are clinically meaningful indicators of muscle quality beyond muscle size alone. The GM contributes substantially to pelvic stability and high-demand locomotor tasks, and altered gluteal muscle composition has been shown to differentiate fallers from non-fallers in older adults^4^. Consistent with this, we observed broad associations between GM fat-related phenotypes and frailty-related traits, and prior work confirms that GM IMAT varies with physical activity^5^.

Interpretation of fat fraction requires caution, as myosteatosis encompasses multiple lipid depots, each with distinct physiological implications^32^. Dixon-derived fat fraction provides a scalable measure^1^ consistent with links between ectopic fat and insulin resistance. CT-based studies demonstrate that myosteatosis relates to hyperinsulinaemia and insulin resistance independent of overall adiposity, and longitudinal evidence links myosteatosis to incident diabetes^33^. Ectopic lipid accumulation frequently coexists with loss of contractile tissue, suggesting that the fat fraction may serve as a surrogate for progressive muscle loss in metabolically compromised individuals, while also capturing local alterations that contribute to reduced strength and performance in T2D^34^.

S2S distances provided spatially resolved insight into regional remodelling beyond what volume and fat fraction capture. Statistical parametric maps revealed distinct patterns of inward and outward deformation associated with anthropometry, lifestyle, and disease, reflecting the GM’s heterogeneous architecture and region-specific functional roles^2,35^. Ageing was characterised by widespread inward deformation consistent with atrophy, whereas BMI was associated with outward deformation indicating region-specific expansion. Higher handgrip strength and vigorous physical activity were associated with outward deformation, consistent with more robust GM morphology in stronger, more active individuals.

Disease-related maps highlighted marked sex differences. In men, T2D was associated with predominantly inward deformation, whereas in women it was mainly associated with outward deformation. A plausible explanation is that T2D-related fay increases in women contribute to apparent GM surface expansion, while in men the dominant signal reflects muscle loss and inward remodelling^1,32^. A plausible explanation is that T2D-related fay increases in women contribute to apparent GM surface expansion, while in men the dominant signal reflects muscle loss and inward remodelling. This interplay between sex, ageing and T2D-related muscle changes aligns with evidence that later life hormonal changes exert sex-specific effects on muscle mass and adiposity^36^. Frailty and osteoporosis showed widespread inward deformation, while CVD, especially in men, showed inward remodelling, suggesting a shared adverse musculoskeletal phenotype.

Bidirectional mediation analyses revealed sex-specific and mechanism-specific patterns. In men, GM muscle volume mediated the relationship between T2D and GM fat fraction, highlighting a bidirectional link between muscle structure and metabolic status. In women, muscle volume did not mediate, suggesting that overall muscle size contributes less to fat fraction variation. PCA-derived shape components further revealed sex-specific patterns, with specific regional PCs reflecting lateral and central-superior GM expansions that mediate parts of the T2D–fat fraction relationship in men, and GM size mediating effects in women. This suggests that in women, T2D-related variation in GM S2S distances may contribute to increased fat fraction, potentially reflecting sex-specific morphological adaptations. These findings align with evidence that body fat distribution and ectopic fat show marked sexual dimorphism in individuals with T2D^8^, and with prior mediation analyses linking physical activity, skeletal muscle measures, and glycaemic control in T2D^37^.

Prediction and survival analyses demonstrated that integrating shape descriptors adds value beyond conventional image-derived phenotypes. In case-control models, GM fat fraction was the strongest single MRI-derived predictor, consistent with the literature linking ectopic lipid accumulation to impaired insulin action and poorer metabolic health^38,39^. Prevalent T2D was associated with substantially higher GM fat fraction reinforcing its clinical relevance as a marker of adverse muscle health. Adding PC scores improved diagnostic performance by capturing regional remodelling not reflected in global size or fat fraction. Notably, the PC describing middle-upper posterior and anterior GM variation was associated with lower future risk of T2D, suggesting a structurally protective phenotype.

Strengths include the large sample size and harmonised imaging/phenotyping framework within the UK Biobank. Limitations include selection bias, limited generalisability beyond predominantly European ancestry^40^, exclusion of participants with missing data, low incidence of future T2D (∼0.5%), and relatively short follow-up (4 years). These factors may reduce the power of time-to-event analyses. Future work should validate findings in independent cohorts, extend follow-up, and evaluate predictive value beyond established risk factors. Incorporating genetic and hormonal data into future analyses may be key to uncovering the mechanisms underlying the GM remodelling patterns identified here.

## Conclusions

In this large population-based imaging study, integrating conventional MRI-derived GM composition with mesh-based morphometry revealed that muscle fat fraction and regional shape remodelling capture complementary aspects of muscle health. GM fat fraction emerged as a robust marker of metabolic dysfunction, while shape patterns captured region-specific adaptations related to ageing, lifestyle, and disease. Longitudinal and mediation analyses demonstrated that structural and metabolic pathways linking GM morphology and T2D differ by sex. Importantly, Incorporating shape PCs improved diagnostic and prognostic performance beyond standard phenotypes, highlighting the value of spatially resolved muscle phenotyping. These findings support a framework in which regional structural shrinkage, fatty replacement, and metabolic dysregulation interact to drive progressive muscle deterioration, and suggest that spatially resolved muscle phenotyping may provide sensitive imaging biomarkers of early muscle loss.

## Ethics approval and consent to participate

The data resources used in this study have approval from ethics committees. Full anonymised images and participants metadata from the UK Biobank cohort was obtained through UK Biobank Access Application number 44584. The UK Biobank has approval from the North West Multi-Centre Research Ethics Committee (REC reference: 11/NW/0382), and obtained written informed consent from all participants prior to the study. All methods were performed in accordance with the relevant guidelines and regulations as presented by the relevant authorities, including the Declaration of Helsinki https://www.ukbiobank.ac.uk/learn-more-about-uk-biobank/about-us/ethics.

## Consent for publication

Not applicable.

## Availability of data and materials

The data that support the findings of this study are available from the UK Biobank (https://www.ukbiobank.ac.uk), but restrictions apply to the availability of these data, which were used under license for the current study, and so are not publicly available. Data are however returned by us to the UK Biobank where they will be fully available on request.

## Author Contributions

J.D.B., E.L.T., D.A., H.R, M.N., C.B-B. and M.T. conceived the study. J.D.B., B.W., E.L.T., D.A. H.R., M.N. and M.T. designed the study. M.T. implemented the methods and performed the data analysis. M.T. defined the disease and physiological condition categories. H.R, D.A. and C.B-B. performed the GM annotations. M.T. performed the image and statistical analysis. J.D.B., E.L.T., D.A., H.R, M.N., C.B-B. and M.T. drafted the manuscript. All authors read and approved the manuscript.

## Data Availability

https://www.ukbiobank.ac.uk

## Acknowledgements

We thank Alex Chekholko for providing technical support that expedited our work. This research has been conducted using the UK Biobank Resource under Application Numbers 44584 and 23889.

