## Supplementary_Material for "Gluteus Maximus Shape Reveals Sex-specific Associations between Morphology and Metabolic Dysfunction"

*Phenotype Definitions*

A summary of the fields corresponding to the variables and the codes corresponding to the considered disease traits are provided in Supplementary Data SD1. Anthropometric measurements including age, weight, height, waist and hip circumferences, hand grip strength (HGS) in the dominant hand, metabolic equivalent task (MET), alcohol intake and smoking status, were acquired at the UK Biobank imaging visit with body mass index (BMI) and waist-to-hip ratio were calculated from these ^1^. For our analysis, we categorised alcohol intake frequency as 1 for “Daily or almost daily” and 0 otherwise and smoking status as 1 for ”Current” and 0 for “Previous” and “Never”. Ethnicity was defined based on the self-reported ethnic background at the initial assessment visit (field: 21000). For the purpose of our analysis we categorised ethnic background as follows: 0 for ”White” and 1 for any other ethnic background (due to small numbers of non-white participants in this dataset (3.1%)). Sex was self-reported and included those recorded by the NHS and those obtained at the initial assessment visit (field: 31).

Biological samples for serum creatinine (field: 30700) were measured in , cystatin C (field: 30720) in , IGF-1 (field: 30770) in , creatinine in urine (field: 30510) in and white blood cell count (field: 30000) in **,** were taken on the initial assessment visit. It should be noted that the UK Biobank initial assessment visit preceded the imaging visit by 9.9 ± 2.4 years.

*Disease Definitions*

Disease/category definitions, including T2D, frailty, osteoporosis, and CVD are previously described in detail ^1,2^.

Briefly, type-2 Diabetes (T2D) was defined based on the International Classification of Diseases 10th edition (ICD10) for type-2 diabetes using E11, self-reported codes 1220 or 1223, respectively ^3^ or taking antidiabetic medication. Antidiabetic medication codes were selected from prescription records from a general practitioner (GP) and the self-reported medication fields 6153 and 6177, reporting as regularly taking “insulin” medication and self-reported treatment field 20003 ^4^ reporting insulin or antidiabetic drugs (BNF section 6.1.1 and 6.1.2) (<https://openprescribing.net/bnf/0601/>). All self-reported medication fields were taken from the UK Biobank imaging visit. Frailty was defined using the criteria adopted by Hanlon et al., ^5^ for use with self-reported UK Biobank questionnaire responses, which require the presence of three out of five indicators, including weight loss, exhaustion, no or only light physical activity in the last four weeks, slow walking speed, and low hand grip strength (HGS)^2^. Additional classifications included pre-frailty (one or two of the five indicators), and not frail (none of the indicators). All frailty indicators were recorded during the UK Biobank imaging visit.

Osteoporosis was defined based on the ICD10 M80-M82 and based on WHO criteria to define osteoporosis from the bone mineral density (BMD) T-score measurements, taken from the UK Biobank imaging visit (spine, femoral neck and hip T-score ≤ −2.5) ^6^. Codes for cardiovascular disease (CVD) were selected based on the general presence of cardiac disease (ICD10: I00-I25, I30-I52) as previously described ^7^. All diseases were defined at the time of the imaging visit and before it.

*Mass Univariate Regression Analysis*

We performed mass univariate regression (MUR) analysis using a refined version of the R package *mutools3D**^8^*. The linear regression model is expressed as follows:

,

where, Y is a × matrix containing subjects from a sample of the population under study and is the number of voxels in the mesh, X is the × p design matrix of p known covariates (including the intercept) and the relevant variables for each subject. X is related to Y by the vector of the estimated regression coefficients . Finally is a × matrix which is independent and identically distributed across the subjects and is assumed to be a zero-mean Gaussian process^9^.

To account for multiple comparisons, we applied the false discovery rate (FDR) procedure using the Benjamini-Hochberg method^10^ to all TFCE-derived p-values for each vertex and each model. The estimated regression coefficients for each relevant covariate and their TFCE-derived p-values, after correction for multiple testing, were then displayed at each vertex in the mesh across the whole 3D ASAT anatomy, providing spatially distributed associations and adjusting the p-values derived from TFCE across all vertices using 1,000 permutations. The estimated regression coefficients for the relevant covariates and their corresponding TFCE-derived p-values, after correcting for multiple testing, were then spatially mapped at each vertex within the 3D ASAT anatomy mesh. This procedure allowed us to uncover associations distributed across the anatomical structure. A 3D mesh construction and phenotype mapping of 3D ASAT thickness is outlined in Supplementary Figures S1 and Figure S3.

Supplementary Tables

|  | **Men**  (N = 300) | **Women**  (N = 300) |
| --- | --- | --- |
| **Age** | 61.57 ± 6.73 | 62.37 ± 6.95 |
| **Weight** | 70.87 ± 6.79 | 60.39 ± 5.57 |
| **Height** | 1.77 ± 0.07 | 1.64 ± 0.06 |
| **BMI** | 22.57 ± 1.40 | 22.40 ± 1.43 |
| **Waist circumference** | 82.96 ± 5.88 | 75.79 ± 6.79 |
| **Hip circumference** | 94.37 ± 4.61 | 94.90 ± 4.96 |
| **WHR** | 0.88 ± 0.05 | 0.80 ± 0.06 |
| **Creatinine** | 79.95 ± 10.29 | 63.61 ± 8.71 |
| **Cystatin C** | 0.86 ± 0.10 | 0.80 ± 0.11 |
| **IGF-1** | 22.69 ± 4.82 | 22.63 ± 5.98 |
| **Creatinine in urine** | 9450.54 ± 5585.73 | 6407.55 ± 4386.37 |
| **WBC count** | 6.15 ± 1.49 | 6.45 ± 1.74 |
| **Vigorous MET** | 10.22 ± 9.14 | 7.61 ± 8.60 |
| **Dominant HGS** | 40.08 ± 7.03 | 25.04 ± 5.29 |
| **Alcohol intake frequency** | 50 (16.7%) | 37 (12.3%) |
| **GM Muscle Left Volume** | 939.40 ± 153.66 | 692.82 ± 96.28 |
| **GM Muscle Right Volume** | 972.66 ± 167.57 | 706.69 ± 101.47 |
| **GM IMAT Left Volume** | 3.75 ± 3.88 | 10.64 ± 12.87 |
| **GM IMAT Right Volume** | 3.30 ± 3.59 | 9.13 ± 12.84 |
| **GM Left Fat Fraction** | 10.24 ± 4.25 | 17.32 ± 7.51 |
| **GM Right Fat Fraction** | 8.61 ± 3.77 | 15.07 ± 7.09 |

**Table S1.** Summary statistics (mean ± standard deviation) for continuous variables and counts (%) for discrete variables of the 300-participant sex-specific cohort for the template construction. Abbreviations: BMI: body mass index; WHR: waist-to-hip ratio, IGF-1: insulin-like growth factor 1; WBC: white blood cell; MET: metabolic equivalent task; GM: gluteus maximus; IMAT: intermuscular adipose tissue.

|  | **Men**  (N = 23,364) | **Women**  (N = 24,670) |
| --- | --- | --- |
| **Caucasian** | 22,567 (96.6%) | 23,881 (96.8%) |
| **Age** | 65.46 ± 7.81 | 64.15 ± 7.61 |
| **Weight** | 83.78 ± 13.41 | 69.02 ± 13.19 |
| **Height** | 1.76 ± 0.07 | 1.63 ± 0.06 |
| **BMI** | 27.03 ± 3.93 | 26.10 ± 4.79 |
| **Waist circumference** | 94.92 ± 10.79 | 83.41 ± 11.97 |
| **Hip circumference** | 100.71 ± 7.40 | 100.92 ± 9.96 |
| **WHR** | 0.94 ± 0.07 | 0.83 ± 0.07 |
| **Creatinine** | 81.36 ± 12.08 | 64.08 ± 9.93 |
| **Cystatin C** | 0.91 ± 0.13 | 0.84 ± 0.12 |
| **IGF-1** | 22.42 ± 5.22 | 21.68 ± 5.68 |
| **Creatinine in urine** | 10,637.83 ± 5,947.42 | 6,889.34 ± 4,755.73 |
| **WBC count** | 6.57 ± 1.89 | 6.60 ± 1.75 |
| **Vigorous MET** | 7.78 ± 8.39 | 6.34 ± 7.88 |
| **Dominant HGS** | 38.14 ± 9.08 | 23.51 ± 6.29 |
| **Alcohol intake frequency** | 4,798 (20.5%) | 3,340 (13.5%) |
| **GM Muscle Left Volume** | 1,011.03 ± 183.32 | 718.56 ± 119.70 |
| **GM Muscle Right Volume** | 1,039.84 ± 193.14 | 737.78 ± 127.03 |
| **GM IMAT Left Volume** | 25.53 ± 50.55 | 24.11 ± 40.37 |
| **GM IMAT Right Volume** | 21.15 ± 43.32 | 20.21 ± 34.06 |
| **GM Left Fat Fraction** | 18.03 ± 8.92 | 21.66 ± 9.38 |
| **GM Right Fat Fraction** | 15.77 ± 8.36 | 19.16 ± 8.87 |

**Table S2.** Summary statistics (mean ± standard deviation) for continuous variables and counts (%) for discrete variables in the full cohort (23,364 men and 24,670 women). Abbreviations: BMI: body mass index; WHR: waist-to-hip ratio; IGF-1: insulin-like growth factor 1; WBC: white blood cell; MET: metabolic equivalent task; HGS: hand grip strength; GM: gluteus maximus; IMAT: intermuscular adipose tissue.GM: gluteus maximus; IMAT: intermuscular adipose tissue.

|  | **GM Muscle Left Volume** | **GM Muscle Right Volume** | **GM Muscle Volume** | **GM IMAT Left Volume** | **GM IMAT Right Volume** | **GM IMAT Volume** | **GM Left Fat Fraction** | **GM Right Fat Fraction** | **GM Fat Fraction** |
| --- | --- | --- | --- | --- | --- | --- | --- | --- | --- |
|  | **Women** | | | | | | | | |
| **Age** | **-0.064 (-0.072, -0.057)*** | **-0.063 (-0.070, -0.056)*** | **-0.063 (-0.071, -0.056)*** | 0.009 (-0.002, 0.021) | 0.010 (-0.001, 0.022) | 0.009 (-0.002, 0.021) | **0.098 (0.085, 0.110)*** | **0.094 (0.082, 0.107)*** | **0.097 (0.084, 0.109)*** |
| **Non-White** | **-0.118 (-0.151, -0.084)*** | **-0.102 (-0.135, -0.069)*** | **-0.108 (-0.141, -0.076)*** | **-0.079 (-0.131, -0.027)*** | **-0.088 (-0.139, -0.036)*** | **-0.079 (-0.131, -0.027)*** | **-0.081 (-0.136, -0.026)*** | **-0.096 (-0.151, -0.042)*** | **-0.089 (-0.144, -0.034)*** |
| **BMI** | **0.230 (0.223, 0.236)*** | **0.256 (0.250, 0.263)*** | **0.246 (0.240, 0.252)*** | **0.390 (0.380, 0.400)*** | **0.377 (0.367, 0.387)*** | **0.390 (0.380, 0.400)*** | **0.432 (0.422, 0.443)*** | **0.430 (0.419, 0.440)*** | **0.434 (0.423, 0.444)*** |
| **WHR** | **0.013 (0.005, 0.022)*** | **0.023 (0.015, 0.031)*** | **0.019 (0.011, 0.027)*** | **0.036 (0.023, 0.049)*** | **0.038 (0.025, 0.051)*** | **0.036 (0.023, 0.049)*** | **0.123 (0.109, 0.136)*** | **0.135 (0.121, 0.149)*** | **0.129 (0.116, 0.143)*** |
| **Creatinine** | **0.116 (0.106, 0.125)*** | **0.111 (0.102, 0.120)*** | **0.115 (0.106, 0.124)*** | **-0.129 (-0.144, -0.115)*** | **-0.126 (-0.140, -0.111)*** | **-0.129 (-0.144, -0.115)*** | **-0.240 (-0.255, -0.224)*** | **-0.241 (-0.256, -0.225)*** | **-0.242 (-0.257, -0.226)*** |
| **Cystatin C** | **-0.043 (-0.051, -0.036)*** | **-0.044 (-0.051, -0.036)*** | **-0.045 (-0.052, -0.037)*** | **0.081 (0.069, 0.093)*** | **0.077 (0.065, 0.089)*** | **0.081 (0.069, 0.093)*** | **0.143 (0.131, 0.156)*** | **0.147 (0.134, 0.160)*** | **0.146 (0.133, 0.159)*** |
| **IGF-1** | **0.032 (0.026, 0.038)*** | **0.029 (0.023, 0.035)*** | **0.031 (0.025, 0.037)*** | **-0.048 (-0.057, -0.038)*** | **-0.045 (-0.054, -0.036)*** | **-0.048 (-0.057, -0.038)*** | **-0.081 (-0.091, -0.072)*** | **-0.080 (-0.090, -0.071)*** | **-0.081 (-0.091, -0.072)*** |
| **Creatinine in urine** | **0.015 (0.008, 0.022)*** | **0.017 (0.009, 0.024)*** | **0.016 (0.009, 0.023)*** | **0.022 (0.011, 0.034)*** | **0.027 (0.015, 0.038)*** | **0.022 (0.011, 0.034)*** | **0.034 (0.022, 0.046)*** | **0.036 (0.024, 0.048)*** | **0.035 (0.024, 0.047)*** |
| **WBC** | **-0.041 (-0.047, -0.035)*** | **-0.039 (-0.045, -0.033)*** | **-0.040 (-0.047, -0.034)*** | **0.011 (0.001, 0.021)*** | 0.009 (-0.001, 0.019) | **0.011 (0.001, 0.021)*** | **0.025 (0.015, 0.035)*** | **0.024 (0.014, 0.034)*** | **0.025 (0.015, 0.035)*** |
| **Time since initial visit** | **-0.006 (-0.013, -0.000)*** | -0.002 (-0.009, 0.004) | -0.005 (-0.011, 0.001) | **0.025 (0.015, 0.035)*** | **0.022 (0.013, 0.032)*** | **0.025 (0.015, 0.035)*** | **0.021 (0.011, 0.031)*** | **0.016 (0.006, 0.026)*** | **0.019 (0.009, 0.029)*** |
| **Vigorous MET** | **0.051 (0.045, 0.057)*** | **0.051 (0.045, 0.057)*** | **0.052 (0.046, 0.058)*** | -0.006 (-0.016, 0.003) | -0.005 (-0.014, 0.005) | -0.006 (-0.016, 0.003) | **-0.045 (-0.055, -0.035)*** | **-0.045 (-0.055, -0.035)*** | **-0.045 (-0.055, -0.035)*** |
| **Dominant HGS** | **0.186 (0.176, 0.197)*** | **0.191 (0.181, 0.202)*** | **0.192 (0.181, 0.202)*** | **0.023 (0.006, 0.039)*** | **0.021 (0.005, 0.037)*** | **0.023 (0.006, 0.039)*** | **-0.072 (-0.089, -0.054)*** | **-0.076 (-0.093, -0.058)*** | **-0.074 (-0.092, -0.057)*** |
| **Alcohol intake** | **0.014 (0.007, 0.020)*** | **0.019 (0.012, 0.025)*** | **0.016 (0.010, 0.022)*** | **0.047 (0.037, 0.057)*** | **0.047 (0.037, 0.057)*** | **0.047 (0.037, 0.057)*** | **0.073 (0.063, 0.084)*** | **0.075 (0.065, 0.086)*** | **0.075 (0.064, 0.085)*** |
| **T2D** | -0.012 (-0.045, 0.021) | 0.003 (-0.030, 0.035) | -0.005 (-0.036, 0.027) | **0.402 (0.352, 0.453)*** | **0.413 (0.362, 0.463)*** | **0.402 (0.352, 0.453)*** | **0.154 (0.100, 0.208)*** | **0.169 (0.115, 0.223)*** | **0.162 (0.109, 0.216)*** |
| **Frailty** | **-0.075 (-0.113, -0.038)*** | **-0.061 (-0.098, -0.024)*** | **-0.068 (-0.104, -0.032)*** | **0.164 (0.106, 0.222)*** | **0.166 (0.109, 0.223)*** | **0.164 (0.106, 0.222)*** | **0.096 (0.035, 0.157)*** | **0.099 (0.037, 0.160)*** | **0.098 (0.037, 0.159)*** |
| **Osteoporosis** | **-0.099 (-0.118, -0.080)*** | **-0.103 (-0.121, -0.084)*** | **-0.103 (-0.121, -0.085)*** | **0.030 (0.001, 0.059)*** | **0.031 (0.002, 0.060)*** | **0.030 (0.001, 0.059)*** | **0.034 (0.003, 0.065)*** | **0.040 (0.009, 0.071)*** | **0.037 (0.006, 0.068)*** |
| **CVD** | **-0.028 (-0.041, -0.014)*** | **-0.024 (-0.037, -0.010)*** | **-0.025 (-0.039, -0.012)*** | **0.032 (0.011, 0.053)*** | **0.032 (0.011, 0.053)*** | **0.032 (0.011, 0.053)*** | **0.062 (0.040, 0.084)*** | **0.068 (0.045, 0.090)*** | **0.065 (0.043, 0.087)*** |
| **Age × T2D** | **-0.037 (-0.069, -0.005)*** | **-0.040 (-0.071, -0.009)*** | **-0.040 (-0.071, -0.009)*** | **-0.096 (-0.145, -0.047)*** | **-0.127 (-0.176, -0.078)*** | **-0.096 (-0.145, -0.047)*** | 0.041 (-0.011, 0.094) | 0.034 (-0.019, 0.086) | 0.038 (-0.014, 0.090) |
|  | **Men** | | | | | | | | |
| **Age** | **-0.081 (-0.092, -0.070)*** | **-0.094 (-0.105, -0.083)*** | **-0.087 (-0.098, -0.077)*** | **0.053 (0.039, 0.067)*** | **0.049 (0.035, 0.064)*** | **0.053 (0.039, 0.067)*** | **0.108 (0.096, 0.119)*** | **0.103 (0.092, 0.114)*** | **0.106 (0.095, 0.117)*** |
| **Non-White** | **-0.304 (-0.354, -0.254)*** | **-0.326 (-0.375, -0.276)*** | **-0.312 (-0.360, -0.264)*** | -0.065 (-0.129, -0.000) | -0.064 (-0.130, 0.001) | -0.065 (-0.129, -0.000) | -0.019 (-0.069, 0.032) | -0.023 (-0.073, 0.028) | -0.021 (-0.071, 0.029) |
| **BMI** | **0.510 (0.497, 0.524)*** | **0.544 (0.531, 0.557)*** | **0.534 (0.522, 0.547)*** | **0.648 (0.631, 0.665)*** | **0.640 (0.622, 0.657)*** | **0.648 (0.631, 0.665)*** | **0.518 (0.505, 0.531)*** | **0.515 (0.502, 0.528)*** | **0.520 (0.507, 0.533)*** |
| **WHR** | **-0.166 (-0.182, -0.150)*** | **-0.163 (-0.178, -0.147)*** | **-0.167 (-0.183, -0.152)*** | **-0.047 (-0.068, -0.027)*** | **-0.051 (-0.072, -0.030)*** | **-0.047 (-0.068, -0.027)*** | **0.118 (0.102, 0.134)*** | **0.117 (0.101, 0.133)*** | **0.118 (0.102, 0.134)*** |
| **Creatinine** | **0.093 (0.081, 0.105)*** | **0.091 (0.079, 0.103)*** | **0.093 (0.082, 0.105)*** | **-0.137 (-0.153, -0.121)*** | **-0.135 (-0.151, -0.119)*** | **-0.137 (-0.153, -0.121)*** | **-0.176 (-0.188, -0.164)*** | **-0.175 (-0.188, -0.163)*** | **-0.177 (-0.189, -0.164)*** |
| **Cystatin C** | **-0.046 (-0.058, -0.035)*** | **-0.040 (-0.051, -0.028)*** | **-0.044 (-0.055, -0.032)*** | **0.102 (0.087, 0.117)*** | **0.104 (0.089, 0.119)*** | **0.102 (0.087, 0.117)*** | **0.118 (0.106, 0.130)*** | **0.117 (0.106, 0.129)*** | **0.118 (0.107, 0.130)*** |
| **IGF-1** | **0.054 (0.044, 0.064)*** | **0.053 (0.043, 0.063)*** | **0.055 (0.045, 0.064)*** | **-0.061 (-0.073, -0.048)*** | **-0.059 (-0.072, -0.046)*** | **-0.061 (-0.073, -0.048)*** | **-0.082 (-0.092, -0.072)*** | **-0.083 (-0.093, -0.073)*** | **-0.083 (-0.093, -0.073)*** |
| **Creatinine in urine** | **0.030 (0.021, 0.039)*** | **0.030 (0.022, 0.039)*** | **0.031 (0.022, 0.039)*** | -0.008 (-0.020, 0.003) | -0.011 (-0.023, 0.001) | -0.008 (-0.020, 0.003) | 0.001 (-0.008, 0.010) | 0.000 (-0.009, 0.009) | 0.001 (-0.008, 0.010) |
| **WBC** | **-0.048 (-0.057, -0.039)*** | **-0.045 (-0.054, -0.036)*** | **-0.048 (-0.057, -0.040)*** | 0.011 (-0.000, 0.022) | 0.009 (-0.003, 0.020) | 0.011 (-0.000, 0.022) | **0.018 (0.009, 0.027)*** | **0.016 (0.007, 0.025)*** | **0.017 (0.008, 0.026)*** |
| **Time since initial visit** | -0.005 (-0.014, 0.005) | 0.008 (-0.002, 0.017) | 0.002 (-0.007, 0.012) | **0.043 (0.031, 0.056)*** | **0.049 (0.036, 0.061)*** | **0.043 (0.031, 0.056)*** | 0.010 (-0.000, 0.019) | **0.014 (0.004, 0.024)*** | **0.012 (0.002, 0.021)*** |
| **Vigorous MET** | **0.080 (0.071, 0.089)*** | **0.078 (0.069, 0.087)*** | **0.080 (0.071, 0.089)*** | **-0.034 (-0.046, -0.022)*** | **-0.033 (-0.044, -0.021)*** | **-0.034 (-0.046, -0.022)*** | **-0.059 (-0.068, -0.050)*** | **-0.056 (-0.066, -0.047)*** | **-0.058 (-0.067, -0.049)*** |
| **Dominant HGS** | **0.186 (0.174, 0.197)*** | **0.176 (0.164, 0.187)*** | **0.183 (0.172, 0.195)*** | **-0.023 (-0.038, -0.008)*** | **-0.024 (-0.039, -0.009)*** | **-0.023 (-0.038, -0.008)*** | **-0.071 (-0.083, -0.060)*** | **-0.073 (-0.085, -0.062)*** | **-0.073 (-0.084, -0.061)*** |
| **Alcohol intake** | **0.020 (0.011, 0.028)*** | **0.027 (0.018, 0.035)*** | **0.023 (0.015, 0.031)*** | **0.022 (0.011, 0.033)*** | **0.020 (0.009, 0.032)*** | **0.022 (0.011, 0.033)*** | **0.040 (0.032, 0.049)*** | **0.040 (0.032, 0.049)*** | **0.041 (0.032, 0.049)*** |
| **T2D** | **-0.144 (-0.181, -0.106)*** | **-0.113 (-0.150, -0.076)*** | **-0.130 (-0.166, -0.094)*** | **0.367 (0.318, 0.415)*** | **0.363 (0.314, 0.412)*** | **0.367 (0.318, 0.415)*** | **0.248 (0.209, 0.286)*** | **0.245 (0.208, 0.283)*** | **0.248 (0.210, 0.286)*** |
| **Frailty** | **-0.180 (-0.246, -0.114)*** | **-0.236 (-0.301, -0.171)*** | **-0.202 (-0.265, -0.139)*** | **0.260 (0.176, 0.345)*** | **0.225 (0.139, 0.311)*** | **0.260 (0.176, 0.345)*** | **0.194 (0.127, 0.260)*** | **0.228 (0.162, 0.294)*** | **0.211 (0.146, 0.277)*** |
| **Osteoporosis** | **-0.223 (-0.267, -0.179)*** | **-0.206 (-0.250, -0.162)*** | **-0.216 (-0.259, -0.174)*** | **0.094 (0.037, 0.151)*** | **0.096 (0.038, 0.154)*** | **0.094 (0.037, 0.151)*** | **0.076 (0.031, 0.121)*** | **0.065 (0.021, 0.110)*** | **0.071 (0.027, 0.116)*** |
| **CVD** | **-0.068 (-0.088, -0.049)*** | **-0.069 (-0.088, -0.050)*** | **-0.071 (-0.089, -0.052)*** | 0.004 (-0.021, 0.029) | 0.011 (-0.015, 0.036) | 0.004 (-0.021, 0.029) | **0.061 (0.042, 0.081)*** | **0.063 (0.044, 0.083)*** | **0.063 (0.043, 0.082)*** |
| **Age × T2D** | -0.027 (-0.063, 0.009) | -0.027 (-0.062, 0.009) | -0.029 (-0.064, 0.005) | 0.003 (-0.043, 0.050) | -0.005 (-0.052, 0.042) | 0.003 (-0.043, 0.050) | **0.038 (0.002, 0.075)*** | **0.039 (0.003, 0.075)*** | **0.039 (0.003, 0.075)*** |

**Table S3.** Summary of regression coefficients for the linear regression models for GM muscle volume, IMAT volume and fat fraction, (left, right, both) representing the associations with relevant baseline characteristics on women and men separately. Values are shown in standardised beta coefficients and 95% confidence interval (CI) values in parentheses. An asterisk (*) indicate statistically significant for FDR-adjusted p-value < 0.05. Significant associations are shown in **bold**. Abbreviations: BMI: body mass index; WHR: waist-to-hip ratio; IGF-1: insulin-like growth factor 1; WBC: white blood cell; MET: metabolic equivalent task; HGS: hand grip strength; T2D: type-2 diabetes; CVD: cardiovascular disease; GM: gluteus maximus; IMAT: intermuscular adipose tissue.

|  | **Men** | | **Women** | |
| --- | --- | --- | --- | --- |
|  | Unstandardised | Unstandardised | Unstandardised | Unstandardised |
|  | **GM Left S2S distances (mm)** | | | |
| **T2D** | −0.38 (0.24, 60.39%) | 0.27 (0.09, 5.33%) | −0.42 (0.25, 8.41%) | 0.42 (0.33, 64.07%) |
| **Frailty** | −0.47 (0.24, 79.61%) | 0.04 (0.05, 0.01%) | −0.37 (0.25, 52.77%) | 0.37 (0.17, 8.32%) |
| **Osteoporosis** | −0.66 (0.33, 93.53%) | - | −0.37 (0.25, 87.23%) | 0.25 (0.12, 4.54%) |
| **CVD** | −0.24 (0.14, 93.62%) | - | −0.12 (0.08, 56.78%) | 0.12 (0.04, 1.69%) |
|  | **GM Right S2S distances (mm)** | | | |
| **T2D** | −0.33 (0.24, 56.56%) | 0.27 (0.09, 6.75%) | −0.45 (0.18, 5.2%) | 0.40 (0.27, 64.52%) |
| **Frailty** | −0.71 (0.33, 87.87%) | 0.05 (0.00, 0.01%) | −0.36 (0.22, 52.81%) | 0.40 (0.13, 7.24%) |
| **Osteoporosis** | −0.66 (0.38, 93.73%) | 0.34 (0.05, 0.12%) | −0.45 (0.27, 87.84%) | 0.27 (0.18, 3.71%) |
| **CVD** | −0.24 (0.09, 89.61%) | - | −0.13 (0.09, 56.38%) | 0.18 (0.09, 4.46%) |

**Table S4.** Significance areas for covariates of the MUR models for the disease states of the model for the left GM and right GM (N=38,868). The total area has been split into areas of positive and negative associations. The unstandardised (raw) regression coefficients () are presented as median (interquartile range - IQR, Significance area (%)) across all vertices of the left and right GM surfaces for the vertices with statistically significant associations. The unstandardised (raw) regression coefficients are based on the standardised coefficients and the median standard deviation of the S2S distances across all vertices (SD for the S2S distances is 4.74 mm (left & right GM) in men and 4.15 mm (left GM), 4.48 mm (right GM) in women. Abbreviations: T2D: type-2 diabetes; CVD: cardiovascular disease; GM: gluteus maximus; S2S: Surface-to-surface.

|  | **GM Muscle Left Volume (ml)** | **GM Muscle Right Volume (ml)** | **GM Muscle Volume (ml)** | **GM IMAT Left Volume (ml)** | **GM IMAT Right Volume (ml)** | **GM IMAT Volume (ml)** | **GM Left Fat Fraction (%)** | **GM Right Fat Fraction (%)** | **GM Fat Fraction (%)** |
| --- | --- | --- | --- | --- | --- | --- | --- | --- | --- |
|  | **Women** | | | | | | | | |
| **Age at baseline** | **-24.25 (-29.91, -18.60)*** | **-24.13 (-29.94, -18.32)*** | **-48.56 (-59.64, -37.48)*** | 0.69 (-0.85, 2.23) | 0.67 (-0.70, 2.05) | 1.38 (-1.70, 4.46) | **1.07 (0.68, 1.46)*** | **1.07 (0.71, 1.44)*** | **2.15 (1.40, 2.90)*** |
| **Race [Non-white]** | -24.58 (-63.05, 13.90) | -35.15 (-74.62, 4.32) | -59.84 (-135.42, 15.74) | -6.49 (-16.99, 4.02) | -5.61 (-14.89, 3.67) | -12.97 (-33.98, 8.04) | -1.33 (-3.99, 1.33) | -1.65 (-4.16, 0.86) | -3.00 (-8.14, 2.13) |
| **Assessment Centre [Newcastle]** | -2.61 (-13.91, 8.70) | 6.48 (-5.12, 18.09) | 4.42 (-17.73, 26.58) | -3.07 (-6.15, 0.01) | -2.30 (-5.04, 0.44) | -6.13 (-12.29, 0.03) | -0.50 (-1.28, 0.28) | -0.43 (-1.16, 0.31) | -0.91 (-2.41, 0.60) |
| **BMI** | **42.04 (38.16, 45.91)*** | **46.35 (42.16, 50.54)*** | **88.77 (82.64, 94.90)*** | **14.74 (13.85, 15.63)*** | **14.24 (13.10, 15.37)*** | **29.49 (27.70, 31.27)*** | **3.58 (3.38, 3.78)*** | **3.63 (3.41, 3.85)*** | **7.12 (6.72, 7.52)*** |
| **WHR** | -0.11 (-2.60, 2.38) | **3.97 (1.15, 6.79)*** | 2.60 (-0.88, 6.07) | 0.29 (-0.22, 0.81) | -0.78 (-1.69, 0.13) | 0.59 (-0.44, 1.62) | 0.02 (-0.09, 0.13) | -0.05 (-0.18, 0.08) | -0.05 (-0.27, 0.18) |
| **Vigorous MET** | 2.19 (0.30, 4.08) | 1.73 (-0.41, 3.87) | **3.18 (0.54, 5.82)*** | 0.02 (-0.37, 0.41) | -0.10 (-0.79, 0.58) | 0.04 (-0.75, 0.82) | **-0.13 (-0.21, -0.04)*** | **-0.15 (-0.25, -0.05)*** | **-0.27 (-0.44, -0.10)*** |
| **Dominant HGS** | **4.14 (1.03, 7.26)*** | **8.34 (4.81, 11.87)*** | **6.47 (2.14, 10.80)*** | -0.45 (-1.10, 0.19) | -0.20 (-1.35, 0.95) | -0.91 (-2.19, 0.38) | -0.12 (-0.26, 0.02) | -0.11 (-0.27, 0.05) | -0.21 (-0.49, 0.07) |
| **Alcohol intake frequency** | **9.28 (3.05, 15.50)*** | 2.45 (-4.57, 9.48) | 9.92 (1.16, 18.68) | 1.24 (-0.06, 2.53) | 1.54 (-0.69, 3.77) | 2.47 (-0.13, 5.07) | 0.18 (-0.10, 0.46) | 0.07 (-0.25, 0.40) | 0.23 (-0.33, 0.79) |
| **T2D** | 27.88 (-6.26, 62.01) | 27.49 (-8.34, 63.31) | 51.47 (-12.10, 115.04) | **19.07 (10.17, 27.98)*** | **16.98 (7.57, 26.38)*** | **38.14 (20.33, 55.95)*** | **2.85 (0.63, 5.06)*** | **2.92 (0.78, 5.06)*** | **5.85 (1.56, 10.14)*** |
| **2nd Visit** | **-11.02 (-12.95, -9.08)*** | **-10.23 (-12.44, -8.02)*** | **-22.20 (-24.86, -19.55)*** | **1.06 (0.66, 1.45)*** | **1.24 (0.49, 1.98)*** | **2.12 (1.32, 2.91)*** | **0.80 (0.72, 0.89)*** | **0.76 (0.66, 0.86)*** | **1.57 (1.40, 1.73)*** |
| **T2D * 2nd Visit** | **-14.27 (-24.21, -4.34)*** | -7.66 (-19.02, 3.69) | **-20.41 (-33.99, -6.82)*** | 1.57 (-0.45, 3.59) | 2.24 (-1.61, 6.09) | 3.14 (-0.90, 7.19) | 0.42 (-0.02, 0.85) | 0.24 (-0.27, 0.75) | 0.65 (-0.21, 1.52) |
|  | **Men** | | | | | | | | |
| **Age at baseline** | **-35.81 (-43.88, -27.73)*** | **-36.45 (-44.87, -28.04)*** | **-74.04 (-89.86, -58.21)*** | **3.50 (1.36, 5.63)*** | **2.43 (0.52, 4.33)*** | **6.99 (2.72, 11.26)*** | **1.80 (1.47, 2.14)*** | **1.59 (1.28, 1.90)*** | **3.39 (2.75, 4.03)*** |
| **Race [Non-white]** | **-74.49 (-128.73, -20.25)*** | **-89.04 (-145.64, -32.44)*** | **-167.13 (-273.93, -60.34)*** | -12.39 (-26.82, 2.03) | -10.14 (-23.01, 2.72) | -24.79 (-53.64, 4.06) | -0.86 (-3.13, 1.40) | -0.67 (-2.77, 1.43) | -1.54 (-5.88, 2.79) |
| **Assessment Centre [Newcastle]** | -8.13 (-24.93, 8.67) | 1.43 (-16.10, 18.96) | -5.58 (-38.63, 27.47) | -1.42 (-5.88, 3.04) | -0.32 (-4.30, 3.67) | -2.84 (-11.77, 6.08) | -0.07 (-0.77, 0.63) | 0.09 (-0.56, 0.74) | 0.04 (-1.30, 1.38) |
| **BMI** | **81.97 (74.93, 89.00)*** | **96.99 (89.92, 104.06)*** | **177.30 (166.04, 188.56)*** | **23.11 (21.66, 24.57)*** | **20.88 (19.52, 22.24)*** | **46.22 (43.31, 49.14)*** | **4.51 (4.27, 4.75)*** | **4.21 (4.00, 4.42)*** | **8.63 (8.19, 9.07)*** |
| **WHR** | -3.08 (-7.89, 1.74) | -1.60 (-6.30, 3.09) | 1.77 (-4.95, 8.50) | -0.23 (-1.09, 0.62) | -0.59 (-1.40, 0.22) | -0.47 (-2.17, 1.23) | **0.19 (0.04, 0.33)*** | **0.17 (0.04, 0.30)*** | **0.35 (0.10, 0.60)*** |
| **Vigorous MET** | **5.24 (2.36, 8.12)*** | **4.35 (1.54, 7.15)*** | **7.88 (3.84, 11.91)*** | -0.55 (-1.07, -0.04) | -0.46 (-0.95, 0.02) | -1.11 (-2.13, -0.08) | **-0.21 (-0.29, -0.12)*** | **-0.17 (-0.25, -0.10)*** | **-0.37 (-0.53, -0.22)*** |
| **Dominant HGS** | **10.59 (6.84, 14.34)*** | 4.09 (0.44, 7.75) | **7.92 (2.65, 13.18)*** | 0.22 (-0.45, 0.89) | 0.28 (-0.35, 0.91) | 0.44 (-0.89, 1.78) | **-0.19 (-0.30, -0.07)*** | -0.07 (-0.17, 0.03) | **-0.25 (-0.45, -0.05)*** |
| **Alcohol intake frequency** | 10.61 (1.27, 19.95) | 0.77 (-8.39, 9.93) | 12.18 (-1.21, 25.56) | 1.54 (-0.16, 3.25) | 1.87 (0.26, 3.49) | 3.09 (-0.32, 6.49) | -0.06 (-0.34, 0.23) | 0.07 (-0.18, 0.32) | 0.00 (-0.51, 0.51) |
| **T2D** | **-73.63 (-112.38, -34.88)*** | **-69.25 (-109.03, -29.47)*** | **-146.87 (-218.42, -75.32)*** | 3.75 (-5.84, 13.33) | 5.02 (-3.60, 13.64) | 7.49 (-11.68, 26.67) | **2.06 (0.54, 3.57)*** | **1.81 (0.41, 3.21)*** | **3.91 (1.04, 6.79)*** |
| **2nd Visit** | **-18.04 (-21.28, -14.80)*** | **-18.15 (-21.28, -15.01)*** | **-36.20 (-40.63, -31.77)*** | **2.14 (1.58, 2.70)*** | **1.80 (1.27, 2.33)*** | **4.28 (3.16, 5.40)*** | **0.83 (0.73, 0.92)*** | **0.77 (0.69, 0.85)*** | **1.60 (1.43, 1.76)*** |
| **T2D * 2nd Visit** | 0.73 (-11.57, 13.03) | -0.05 (-11.96, 11.86) | -1.03 (-17.80, 15.75) | **2.55 (0.43, 4.67)*** | 1.84 (-0.18, 3.86) | **5.10 (0.86, 9.34)*** | 0.20 (-0.16, 0.55) | 0.26 (-0.05, 0.58) | 0.46 (-0.18, 1.09) |

**Table S5.** Summary of regression coefficients for the linear mixed-effect models for GM muscle volume, IMAT volume, and fat fraction, (left, right, both) representing the associations with relevant baseline characteristics on women and men separately. Values are shown in standardised beta coefficients and 95% confidence interval (CI) values in parentheses. An asterisk (*) indicates statistically significant for FDR-adjusted p-value < 0.05. Significant associations are shown in **bold**. Abbreviations: BMI: body mass index; WHR: waist-to-hip ratio; MET: metabolic equivalent task; HGS: hand grip strength; T2D: type-2 diabetes; CVD: cardiovascular disease; GM: gluteus maximus; IMAT: intermuscular adipose tissue.

| **Left GM** | **Men** | | **Women** | |
| --- | --- | --- | --- | --- |
|  | Median standardised beta coefficients (IQR, Significance area (%)) | | | |
| **Age at baseline** | -0.43 (0.32, 72.83%) | 0.26 (0.13, 3.56%) | -0.42 (0.38, 61.06%) | 0.27 (0.11, 4.06%) |
| **Race [Non-white]** | -2.67 (1.06, 19.84%) | - | -3.10 (1.38, 3.33%) | 1.38 (0.17, 0.42%) |
| **Assessment Centre [Newcastle]** | -0.67 (0.25, 16.95%) | 0.63 (0.31, 7.97%) | -0.62 (0.38, 27.47%) | 0.77 (0.48, 9.46%) |
| **BMI** | -0.38 (0.09, 0.9%) | 1.37 (0.73, 95.22%) | -0.36 (0.17, 1.51%) | 0.99 (0.64, 92.42%) |
| **WHR** | -0.29 (0.20, 39.24%) | - | -0.23 (0.09, 8.78%) | 0.21 (0.09, 23.62%) |
| **Vigorous MET** | - | 0.21 (0.10, 38.76%) | -0.16 (0.03, 0.21%) | 0.18 (0.10, 12.52%) |
| **Dominant HGS** | - | 0.26 (0.16, 81.83%) | - | 0.19 (0.14, 64.92%) |
| **Alcohol intake frequency** | - | - | - | - |
| **T2D** | -1.74 (0.81, 9.66%) | - | - | - |
| **2nd Visit** | -0.49 (0.54, 41.48%) | 0.30 (0.22, 32.58%) | -0.45 (0.41, 41.42%) | 0.30 (0.24, 32.67%) |
| **T2D * 2nd Visit** | - | - | - | - |

**Table S5.** Significance areas for covariates of the mass univariate linear mixed-effect models for the anthropometric variables of the left GM S2S distances (mm) by gender (1,362 men and 1,358 women). The total area has been split into positive and negative association areas. The standardised regression coefficients () are presented as medians (interquartile ranges - IQR), and the significance areas are expressed as percentages (%) of the vertices with statistically significant associations. Abbreviations: BMI: body mass index, WHR: waist-to-hip ratio; MET: metabolic equivalent task; HGS: hand grip strength; T2D: type-2 diabetes; GM: gluteus maximus; IMAT: intermuscular adipose tissue.T2D: type-2 diabetes; GM: gluteus maximus.

| **Right GM** | **Men** | | **Women** | |
| --- | --- | --- | --- | --- |
|  | Median standardised beta coefficients (IQR, Significance area (%)) | | | |
| **Age at baseline** | -0.46 (0.33, 71.87%) | 0.31 (0.11, 2.58%) | -0.46 (0.36, 56.43%) | 0.30 (0.11, 3.76%) |
| **Race [Non-white]** | -2.56 (1.06, 28.38%) | - | -2.56 (1.06, 28.38%) | - |
| **Assessment Centre [Newcastle]** | -0.66 (0.32, 12.61%) | 0.69 (0.39, 22.44%) | -0.74 (0.41, 24.47%) | 0.68 (0.47, 27.65%) |
| **BMI** | -0.37 (0.12, 0.46%) | 1.47 (0.75, 96.92%) | -0.40 (0.18, 0.96%) | 1.05 (0.62, 93.74%) |
| **WHR** | -0.28 (0.14, 40.64%) | - | -0.17 (0.06, 3.47%) | 0.22 (0.11, 19.61%) |
| **Vigorous MET** | - | 0.23 (0.14, 38.83%) | - | 0.18 (0.06, 6.3%) |
| **Dominant HGS** | - | 0.28 (0.18, 59.2%) | -0.11 (0.01, 0.4%) | 0.24 (0.19, 72.1%) |
| **Alcohol intake frequency** | - | - | - | - |
| **T2D** | - | - | - | - |
| **2nd Visit** | -0.51 (0.50, 42.73%) | 0.37 (0.34, 28.37%) | -0.47 (0.48, 38.61%) | 0.30 (0.20, 31.57%) |
| **T2D * 2nd Visit** | - | - | - | - |

**Table S6.** Significance areas for covariates of the mass univariate linear mixed-effect models for the anthropometric variables of the right GM S2S distances (mm) by gender (1,362 men and 1,358 women). The total area has been split into positive and negative association areas. The standardised regression coefficients () are presented as median (interquartile ranges - IQR), and the significance areas as a percentage (%) of the vertices with statistically significant associations. All continuous variables used as fixed effects were standardised before analysis. Abbreviations: BMI: body mass index, WHR: waist-to-hip ratio; MET: metabolic equivalent task; HGS: hand grip strength; T2D: type-2 diabetes; GM: gluteus maximus.

Supplementary Figures

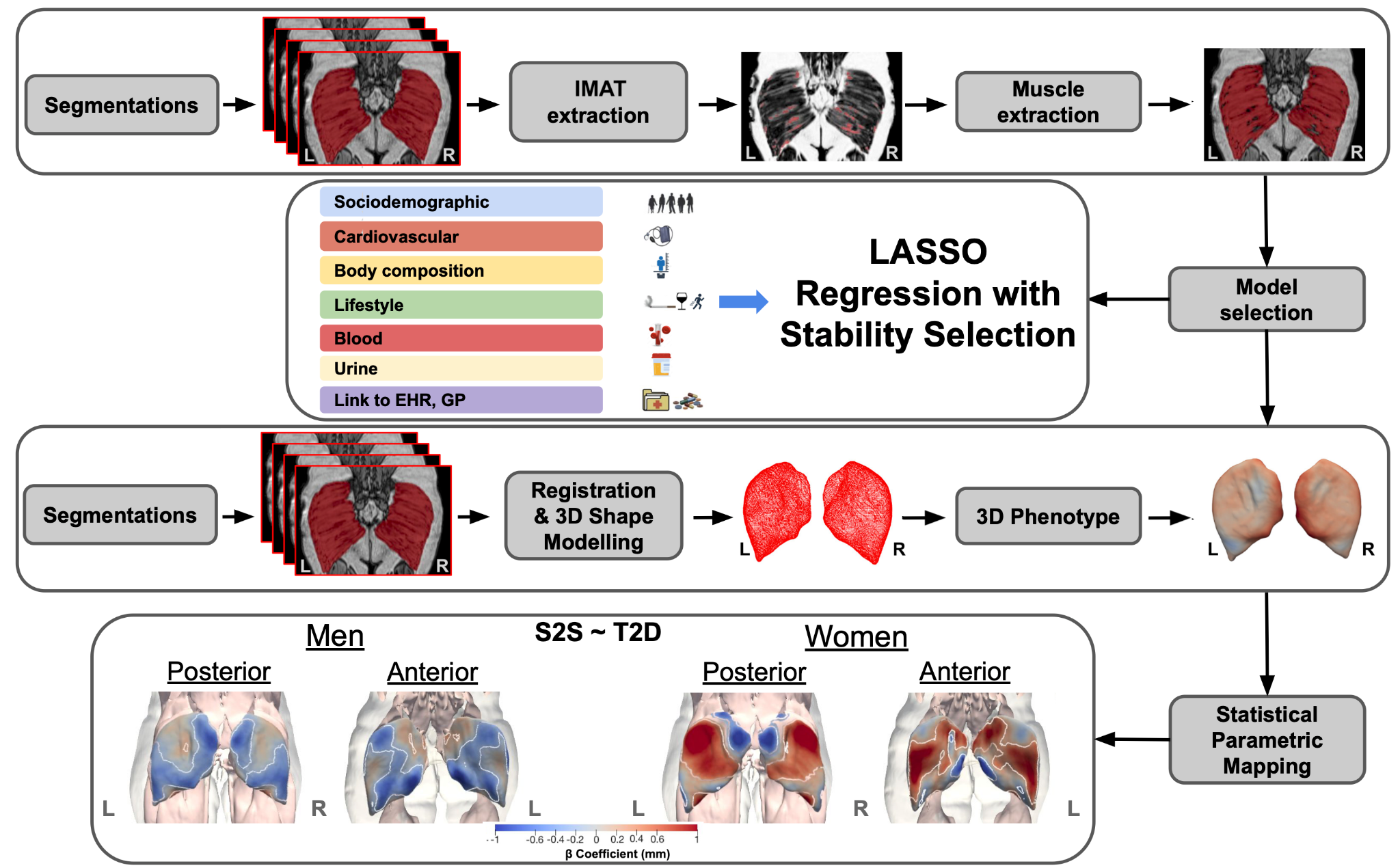

**Figure S1.** Example of GM segmentation on T1 Dixon sequences showing the extraction of IMAT (thresholded at 0.5 fat fraction cut-off) mapped on the fat images and muscle mapped on the opposed phase images. Relevant GM predictors were identified via LASSO with stability selection. Participant segmentations were used to register images, create 3D GM reference meshes for men and women, and compute 3D GM S2S. Associations between S2S and T2D were mapped onto the template meshes. Projections showing the anterior and posterior views for both left (L) and right (R) GM. White contour lines indicate the boundary between statistically significant regions (p < 0.05) after correction for multiple testing, with positive associations in bright red and negative associations in bright blue. Reproduced by kind permission of UK Biobank ©. Abbreviations: LASSO: least absolute shrinkage and selection operator; GM: Gluteus maximus; S2S: Surface-to-surface; T2D: type-2 diabetes; IMAT: intermuscular adipose tissue.

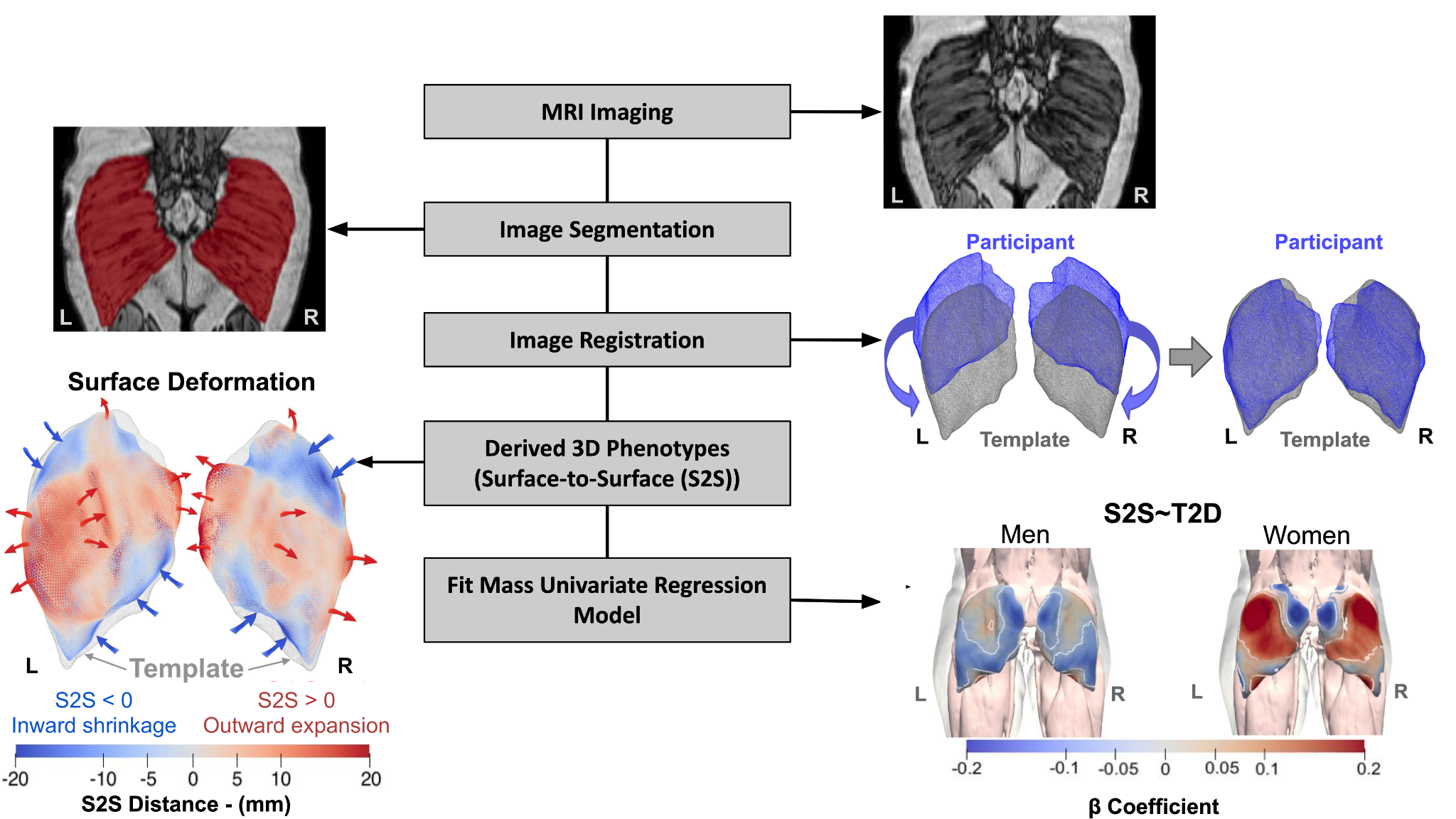
**Figure S2.** Dixon MRI images from UK Biobank are used to segment the GM. GM participant’s segmentation are used to register images to a common space, and create a 3D reference GM template mesh for men and women. The templates were then propagated to each participant's mesh, ensuring anatomical consistency across all participants. 3D GM S2S is computed, mapped to meshes, and analysed via mass univariate regression, with the results of the associations between S2S and T2D visualised on the template mesh for men and women. Projections showing the posterior views for both left (L) and right (R) GM. White contour lines in the MUR model indicate the boundary between statistically significant regions (p < 0.05) after correction for multiple testing, with positive associations in bright red and negative associations in bright blue.Reproduced by kind permission of UK Biobank ©. Abbreviations: GM: Gluteus maximus; S2S: Surface-to-surface; T2D: type-2 diabetes; MUR: Mass univariate regression.

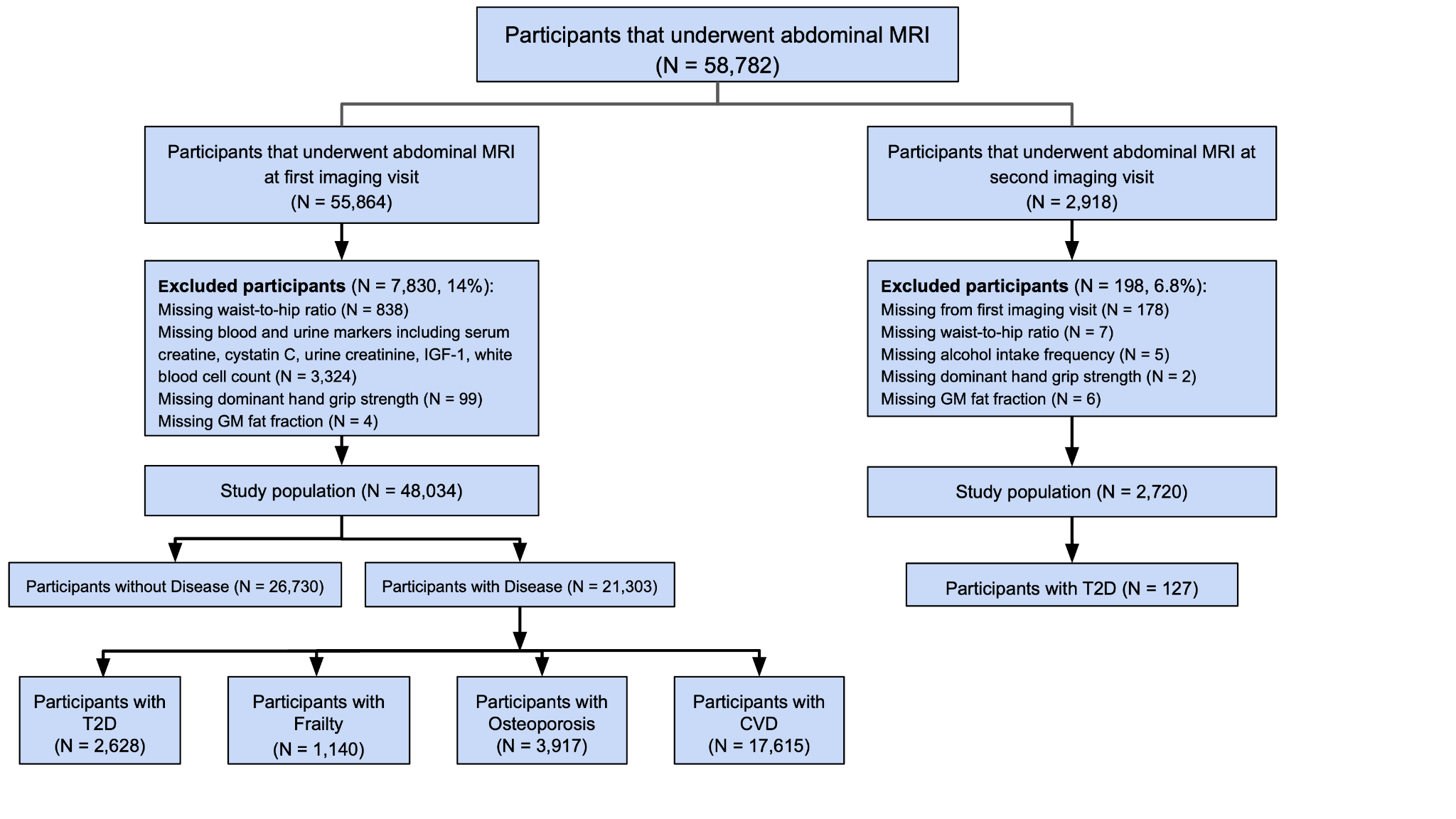

**Figure S3.** Flow diagram of the study population used in this study (N = 48,034; 23,364 men and 24,670 women) of which 2,628 with T2D, 1,140 with frailty, 3,917 with osteoporosis and 17,615 with CVD. Abbreviations: T2D: type-2 diabetes; CVD: cardiovascular disease.

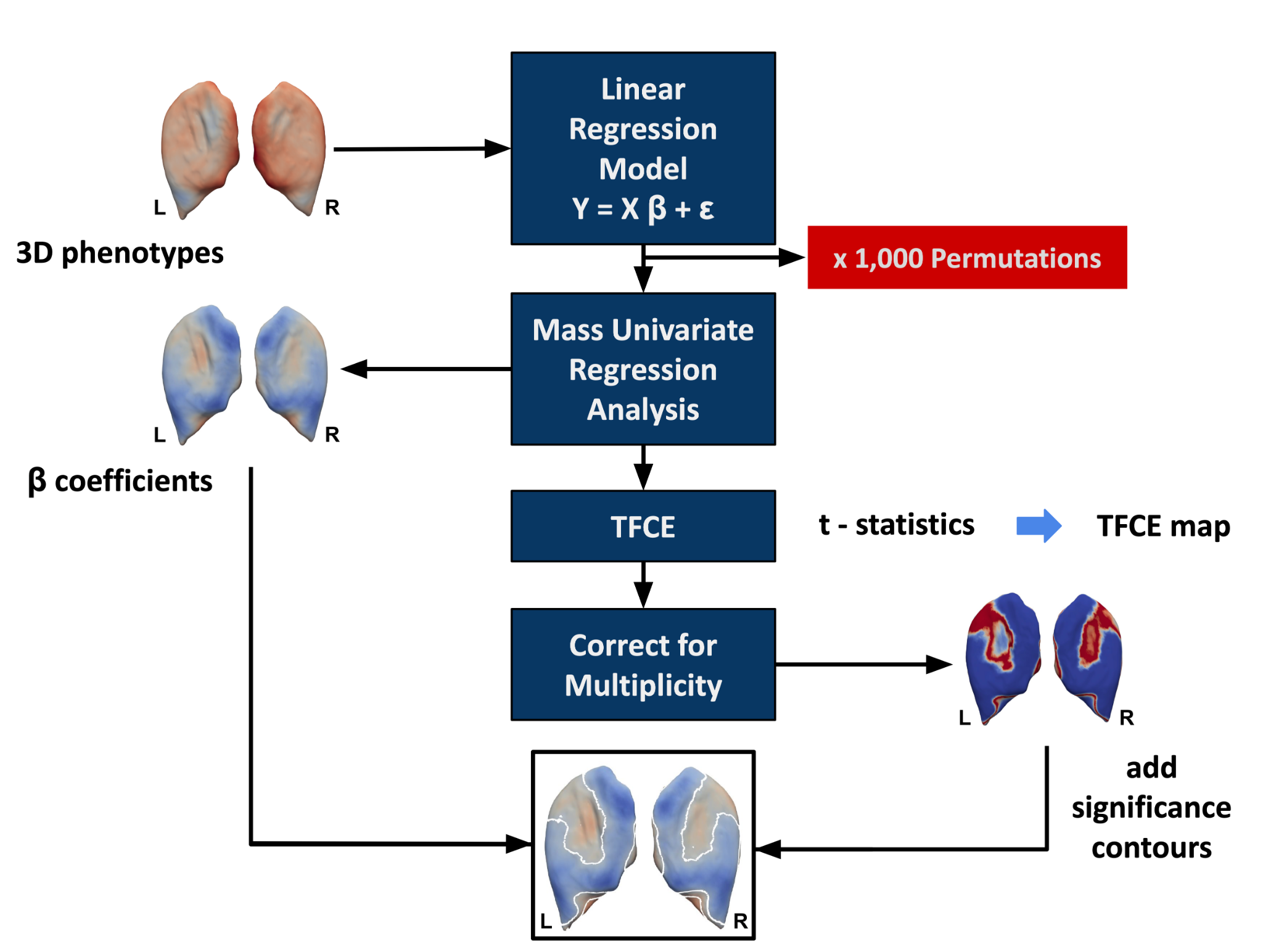
**Figure S4.** Flow diagram for the MUR analysis of the 3D GM S2S phenotypes mapped onto a template surface. The 3D phenotypes are used to construct the linear regression model. MUR analysis produces parameter estimates () and their null distribution via permutation. TFCE is applied to the -statistics from the regression analysis to produce a significance threshold. The associated TFCE-derived -values are corrected for multiple comparisons and mapped onto the GM template mesh for visualisation. This diagram was modified from Biffi et al.^8^. Abbreviations: MUR: Mass univariate regression; TFCE: threshold-free cluster enhancement

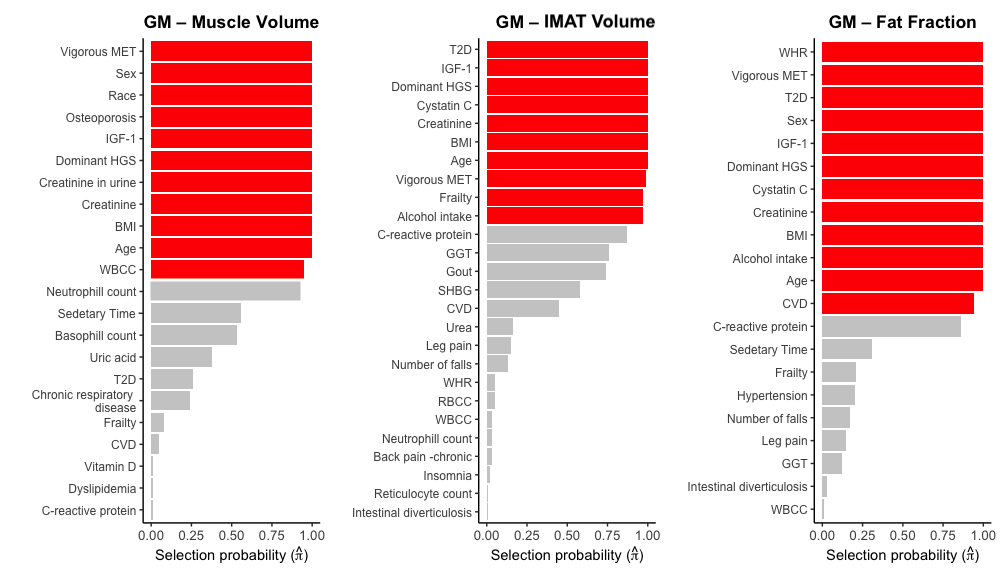

**Figure S5.** Plot showing the covariates selected after stability selection as predictors of GM muscle, IMAT volume and fat fraction. Red bars indicate variables selected after stability selection. Abbreviations: GM: gluteus maximus; IMAT: intermuscular adipose tissue.

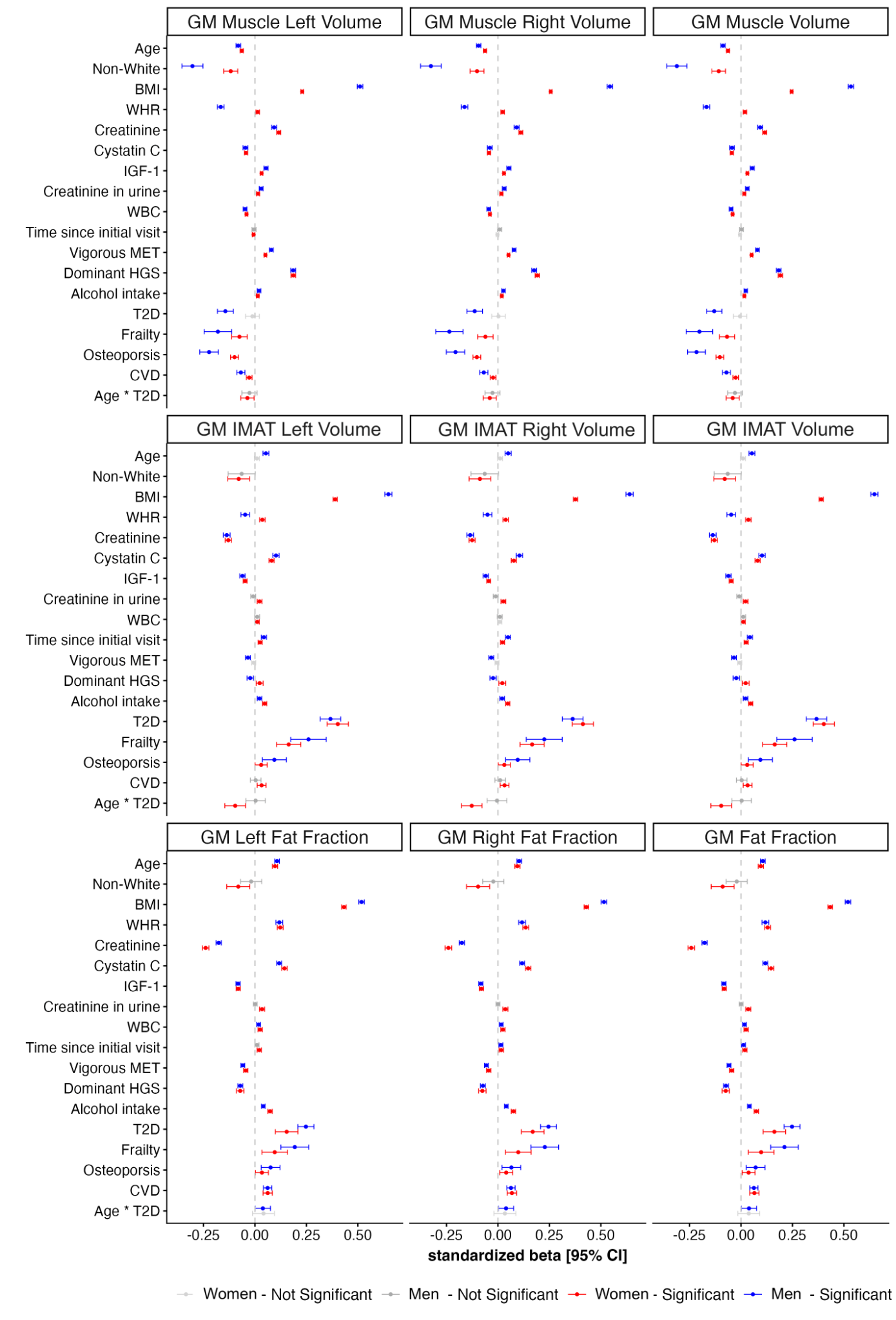

**Figure S6.** Summary of linear regression coefficients for GM muscle, IMAT volume and fat fraction, for left, right and overall GM, representing the associations with the most relevant baseline characteristics in women (N=24,670) and men (N=23,364), separately. Standardised beta coefficients are displayed with 95% confidence intervals. Significant associations for FDR-adjusted p-value < 0.05 are shown in red for women and blue for men, and non-significant associations in grey. Abbreviations: BMI: body mass index; WHR: waist-to-hip ratio, IGF-1: insulin-like growth factor 1; WBC: white blood cell; MET: metabolic equivalent task; T2D: type-2 diabetes; CVD: cardiovascular disease; GM: gluteus maximus; IMAT: intermuscular adipose tissue; FDR: false discovery rate.

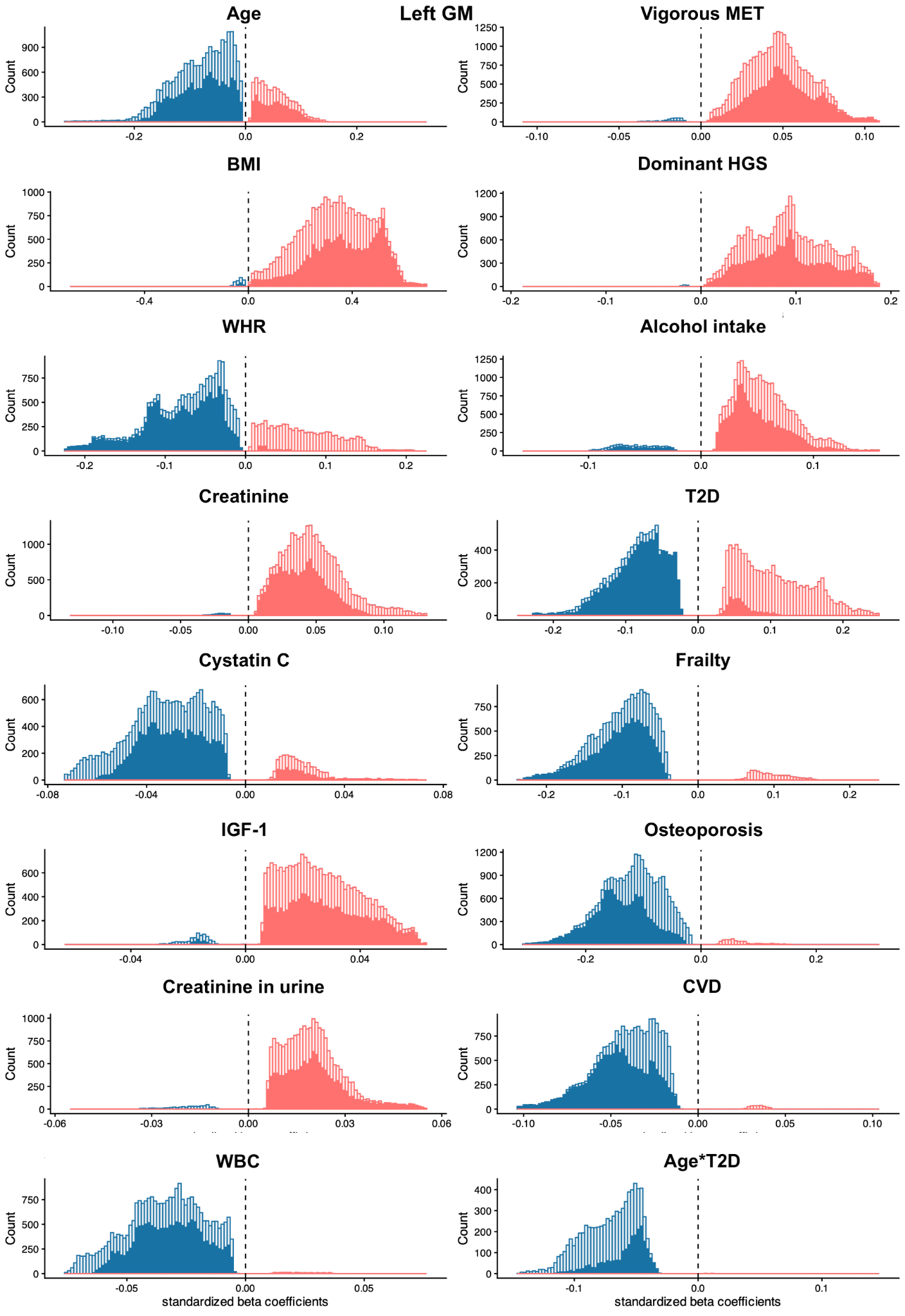

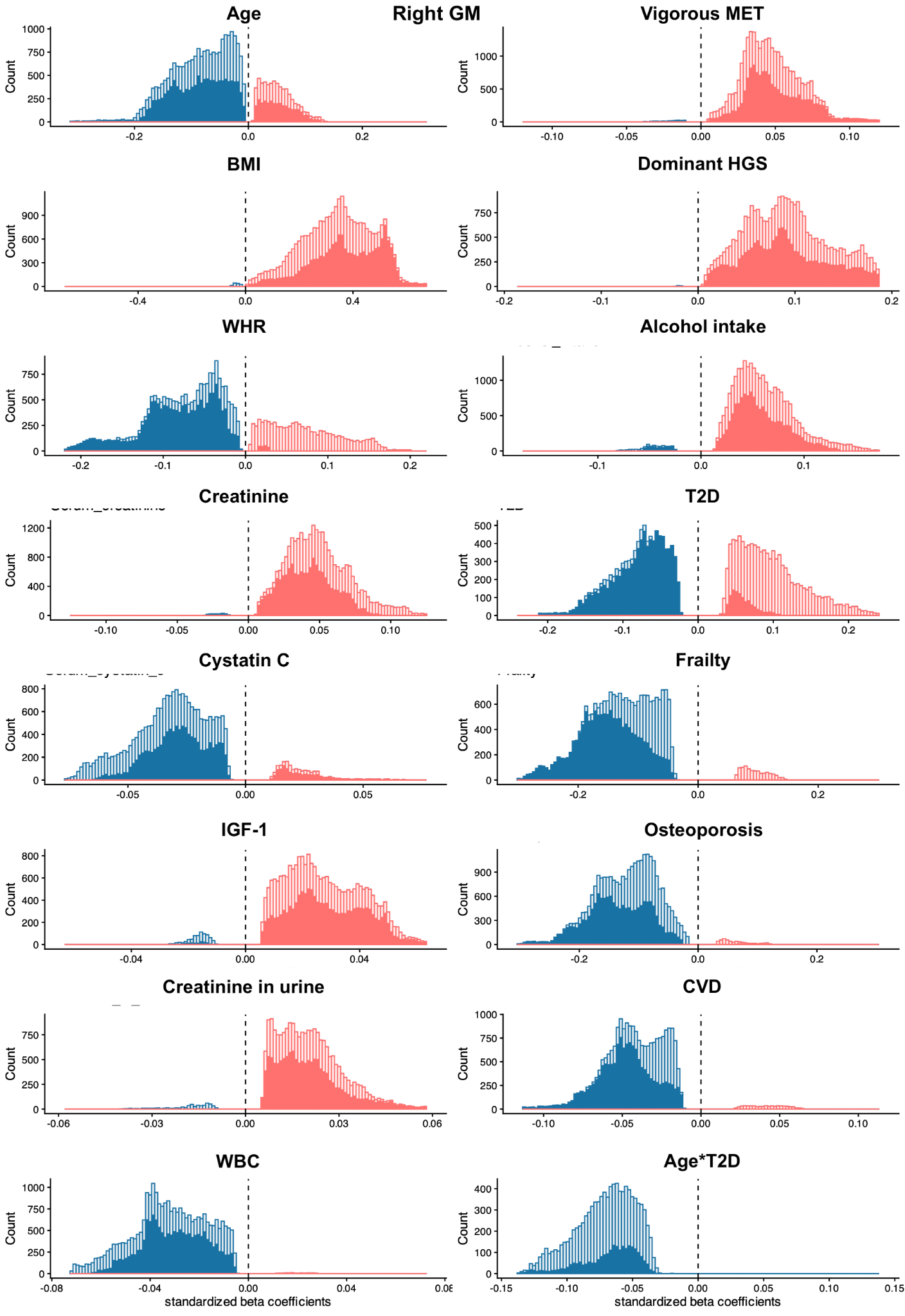
**=**

**Figure S7.** Histograms of the statistically significant regression coefficients across the vertices (approximately 16,000 in men and 13,000 in women) of the GM for each covariate in the model the separated by gender, for each left and right GM. Positive associations are in red and negative associations in blue, with men participants (N=23,364) in a darker colour and women participants (N=24,670) in a lighter colour. The regression (beta) coefficients are provided with units in standard deviations for each covariate. Abbreviations: BMI: body mass index; WHR: waist-to-hip ratio, IGF-1: insulin-like growth factor 1; WBC: white blood cell; MET: metabolic equivalent task; T2D: type-2 diabetes; CVD: cardiovascular disease; GM: gluteus maximus.

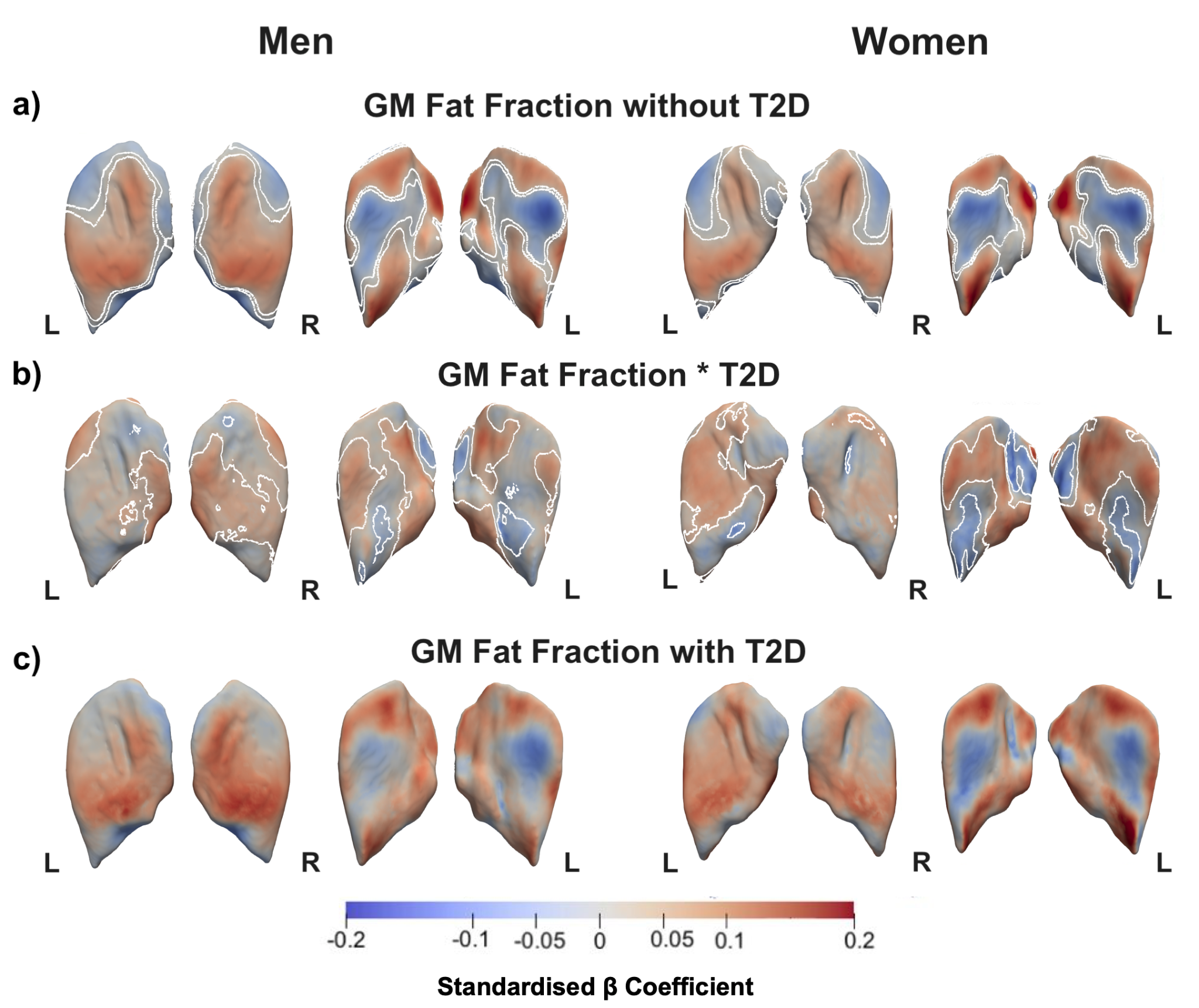

**Figure S8.** Three-dimensional sex-specific SPMs of GM morphology, projections are posterior and anterior views for both left (L) and right (R) GM in both posterior (left plots) and anterior (right plots) views. The SPMs show the local rate of change as a function of GM fat fraction for S2S distances in participants **a)** without T2D and **b)** interactions between GM FF and T2D, **ic)** participants with T2D, referred to the sums of the regression coefficients for GM fat fraction and GM fat fraction*T2D, in men (N=23,364) and women (N=24,670). Positive associations are in red and negative associations are in blue. Regression coefficients () are shown with units in standard deviations for each covariate. Abbreviations: GM: gluteus maximus; T2D: type-2 diabetes; SPMs: statistical parametric maps; S2S: surface-to-surface.

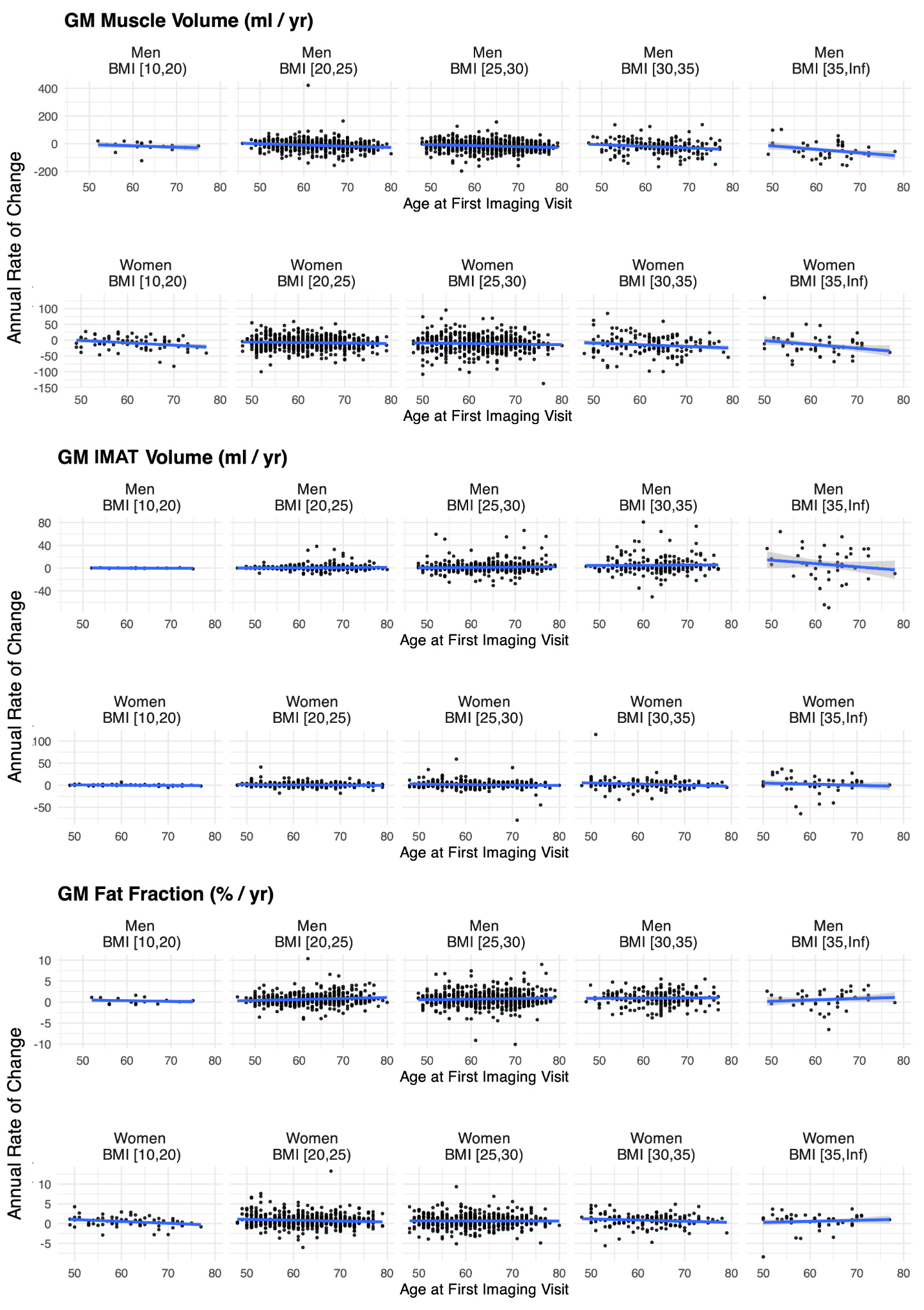

**Figure S9.** Annual changes in gluteus maximus (GM) muscle volume, IMAT volume, and fat fraction between the first and second imaging visits, shown in relation to age at the first imaging visit across BMI categories (10–20, 20–25, 25–30, 30–35, ≥35 kg/m²). Results are presented separately for men (N = 1,362) and women (N = 1,358). Abbreviations: BMI: body mass index; GM: gluteus maximus; IMAT: intermuscular adipose tissue.

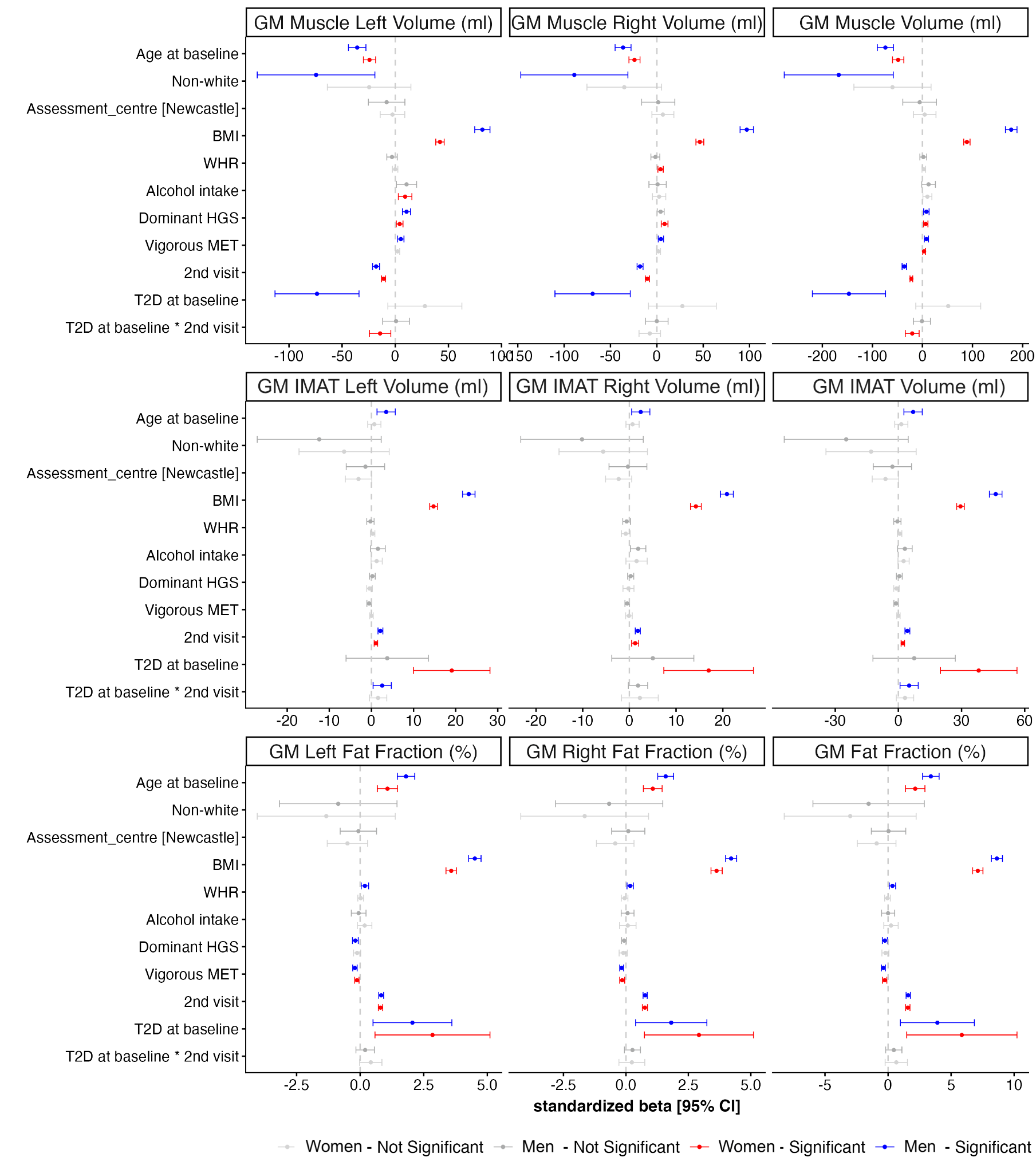

**Figure S10.** Summary of linear mixed effects regression coefficients for GM muscle, IMAT volume and fat fraction, for left, right and overall GM, representing the associations with the most relevant baseline characteristics for men (N=1,362) and women (N=1,358), separately. Standardised beta coefficients are displayed with 95% confidence intervals. Significant associations for FDR-adjusted p-value < 0.05 are shown in red for women and blue for men, and non-significant associations in grey. Abbreviations: BMI: body mass index; WHR: waist-to-hip ratio, MET: metabolic equivalent task; T2D: type-2 diabetes; GM: gluteus maximus; FDR: false discovery rate; IMAT: intermuscular adipose tissue.

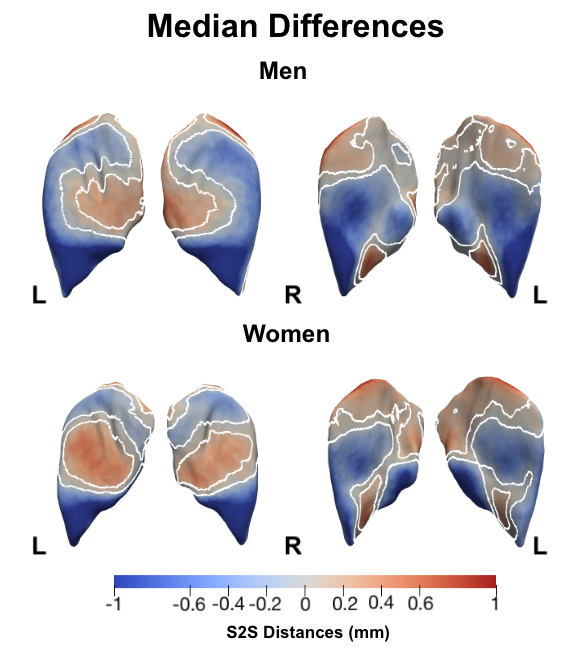
**Figure S11.** Three-dimensional GM morphology, showing the median changes in the GM S2S between the first and second imaging visit for men (N=1,362) and women (N=1,358). Projections are posterior and anterior views for both left (L) and right (R) GM in both posterior (left plots) and anterior (right plots) views. White contour lines indicate the boundary between statistically significant regions (FDR-adjusted p < 0.05) after multiple-testing correction, with positive changes shown in bright red and negative changes in bright blue. Abbreviations: GM: gluteus maximus; S2S: surface-to-surface.

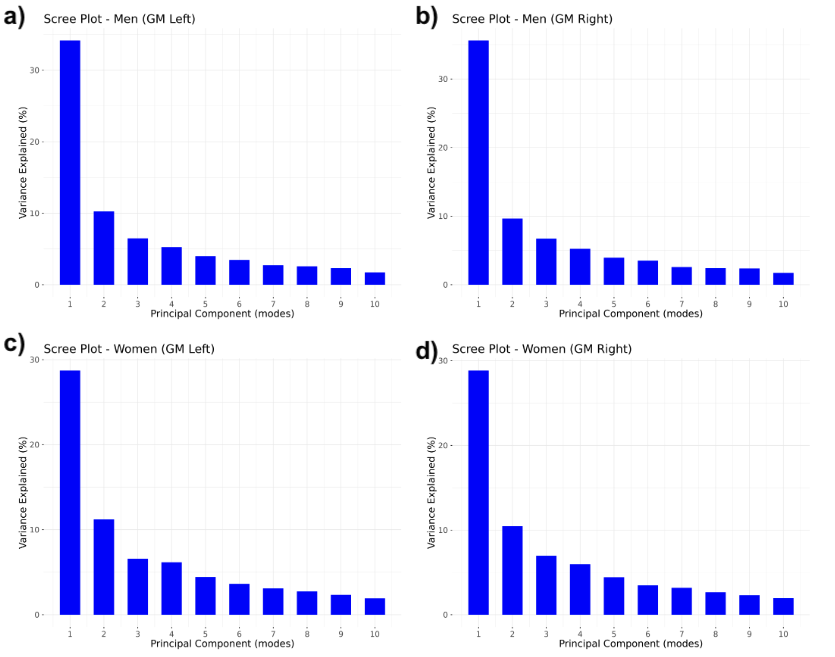
**Figure S12.** The percentage of shape variation explained by the first ten modes of PCA for the **a, b)** left and right GM in men (N=23,364) and **c, d)** women (N=24,670). Abbreviations: PCA: principal component analysis; GM: gluteus maximus.

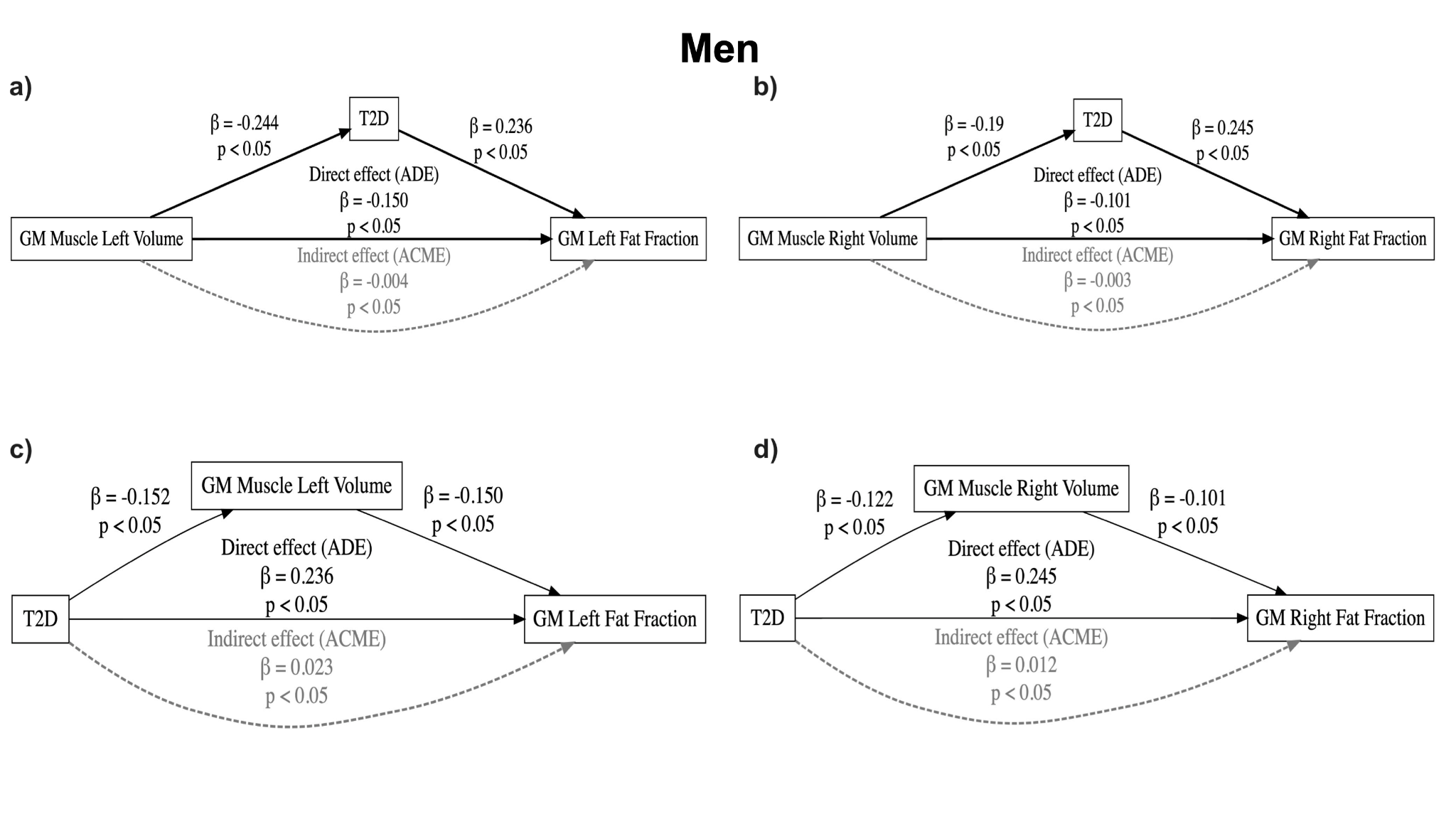
**Figure S13.** Causal mediation analysis on the relationship between GM muscle volume, T2D and GM fat fraction in men (N=23,364). Upper panels **a)** and **b)** show exposure GM left and right muscle volume with mediator T2D. Lower panels **c)** and **d)** show exposure T2D with mediator GM left and right muscle volume. Direct effects are indicated by solid black arrows, whereas mediation effects are indicated by dashed grey arrows. Labels indicate effect size and p-values. Abbreviations: T2D: type-2 diabetes; GM: gluteus maximus.

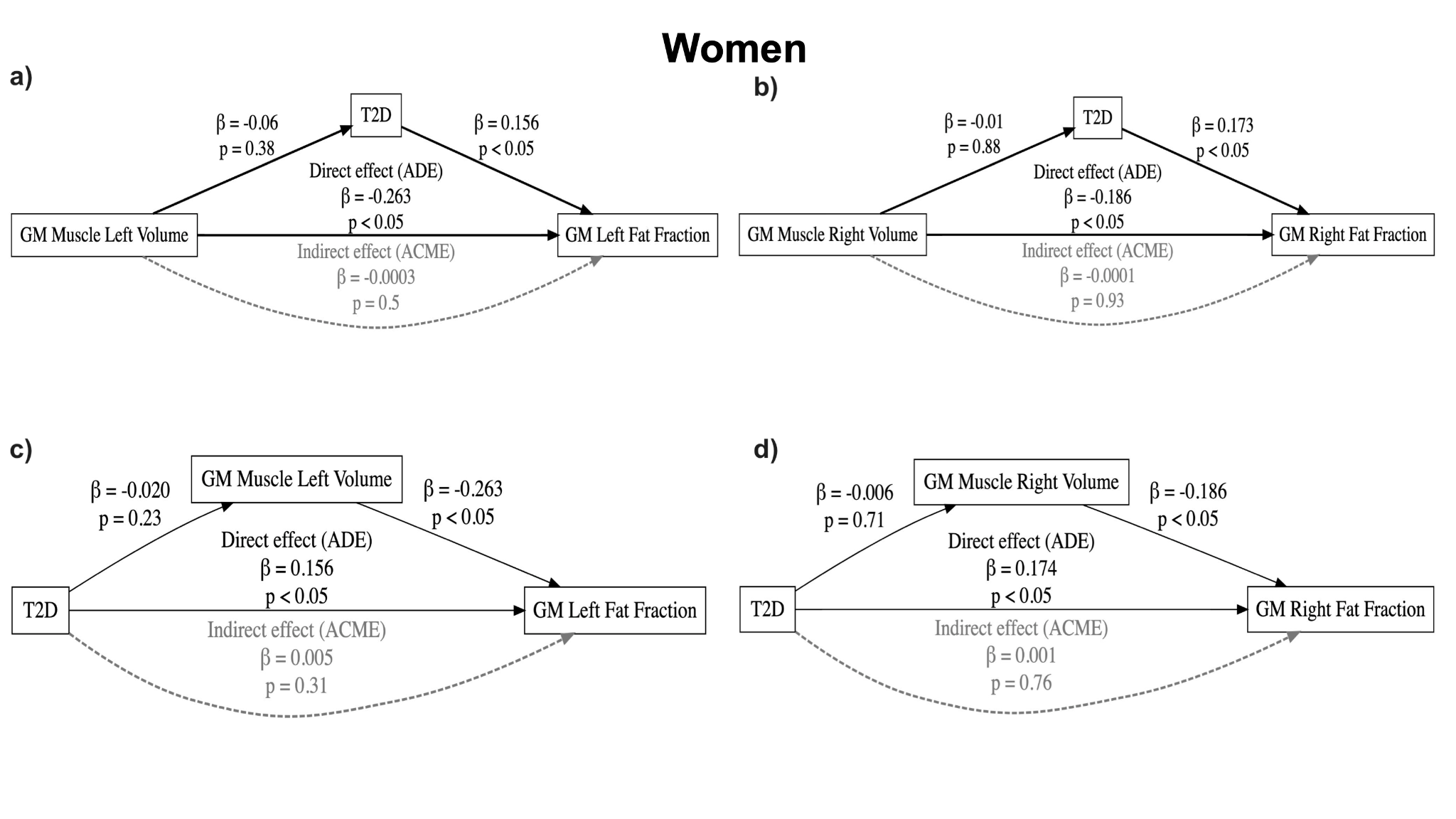
**Figure S14.** Causal mediation analysis on the relationship between GM muscle volume, T2D and GM fat fraction in women (N=24,670). Upper panels **a)** and **b)** show exposure GM left and right muscle volume with mediator T2D. Lower panels **c)** and **d)** show exposure T2D with mediator GM left and right muscle volume. Direct effects are indicated by solid black arrows, whereas mediation effects are indicated by dashed grey arrows. Labels indicate effect size and p-values. Abbreviations: T2D: type-2 diabetes; GM: gluteus maximus.

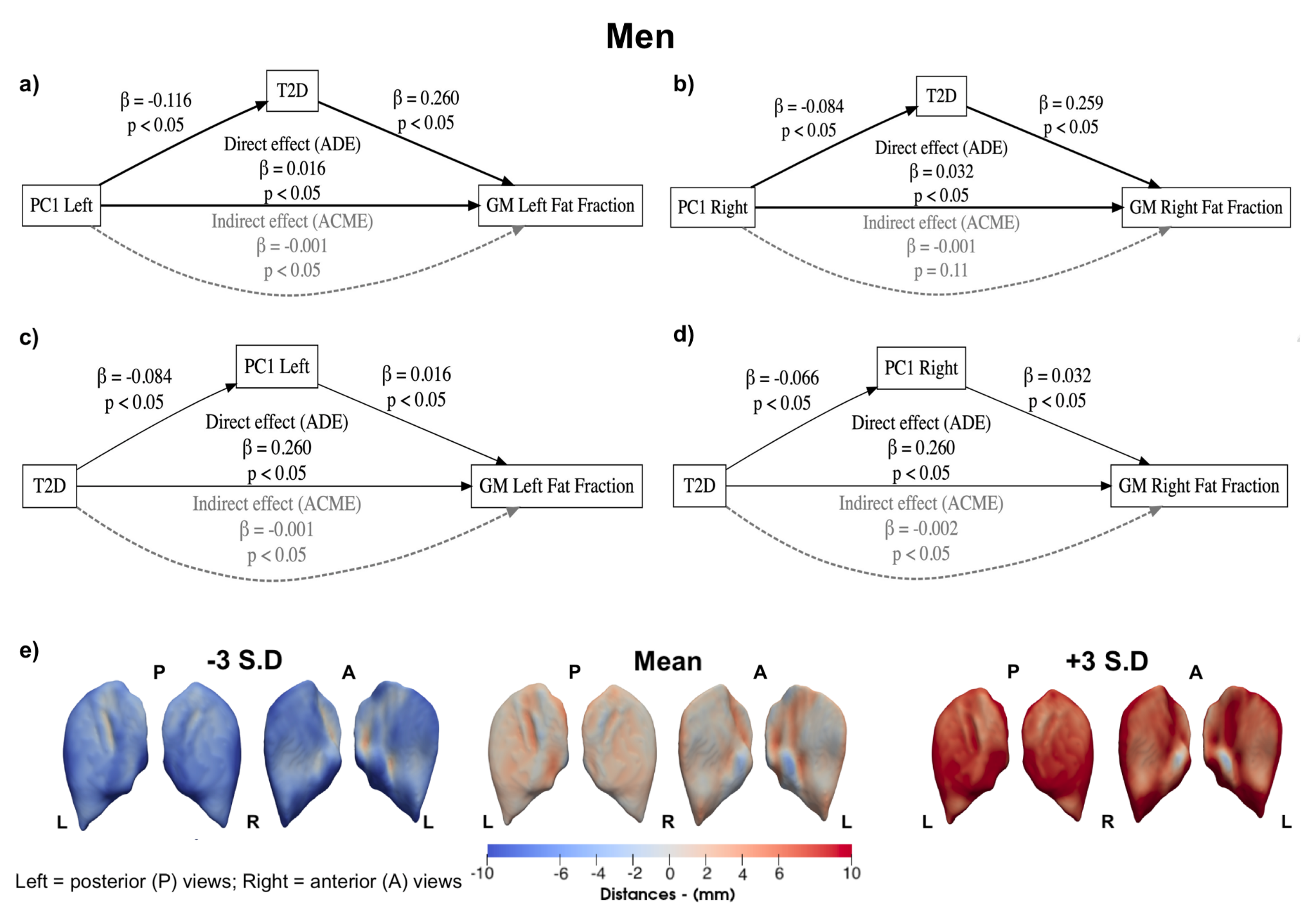

**Figure S15.** Causal mediation analysis on the relationship between PC1 explaining GM size (from smaller to larger), T2D and GM fat fraction in men (N=23,364). Upper panels **a)** and **b)** show exposure PC1 of GM left and right with mediator T2D. Lower panels **c)** and **d)** show exposure T2D with mediator PC1 of GM left and right. **e)** The mean shape and the shape at the ±3 SD are displayed for PC1, showing the S2S distance variation in mm. The GM shape variations are shown in the anterior and posterior views of each GM. Direct effects are indicated by solid black arrows, whereas mediation effects are indicated by dashed grey arrows. Labels indicate effect size and p-values. Abbreviations: PC: principal component; GM: gluteus maximus; S2S: surface-to-surface; T2D: type-2 diabetes.

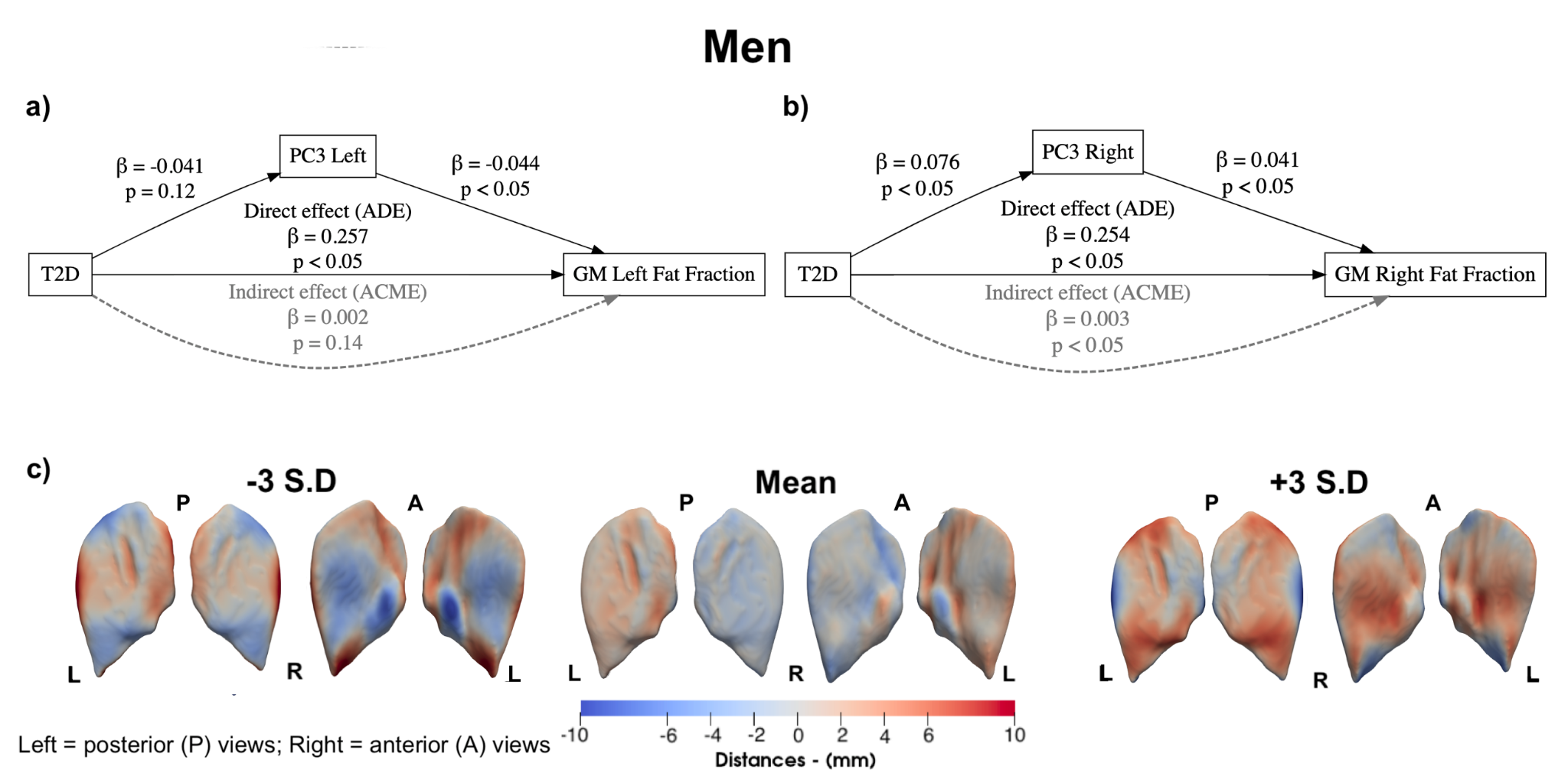

**Figure S16.** Causal mediation analysis on the relationship between PC3, explaining the right GM lateral and iliosacral expansion and shrinkage in the supero-inferior and femoral head regions, T2D and GM fat fraction in men (N=23,364). Upper **a)** and **b)** show exposure T2D with mediator PC3 of GM left and right. T2D did not significantly mediate the effect of PC3 of GM (left and right) on GM fat fraction (left and right). **c)** The mean shape and the shape at the ±3 SD are displayed for PC3, showing the S2S distance variation in mm. The GM shape variations are shown in the anterior and posterior views of each GM. Direct effects are indicated by solid black arrows, whereas mediation effects are indicated by dashed grey arrows. Labels indicate effect size and p-values. Abbreviations: PC: principal component; GM: gluteus maximus; S2S: surface-to-surface; T2D: type-2 diabetes.

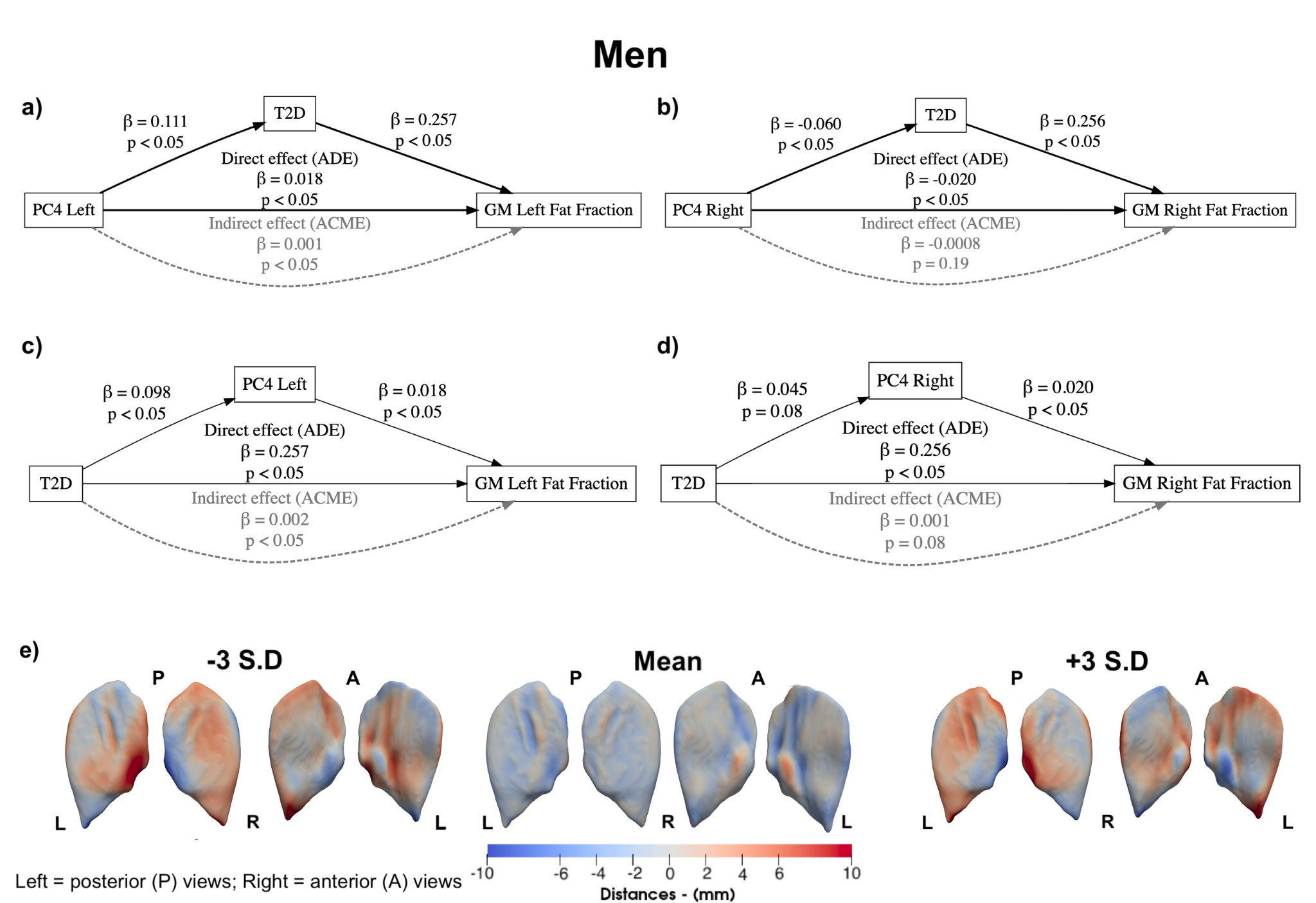

**Figure S17.** Causal mediation analysis on the relationship between PC4, explaining the left GM central/superior/inferior expansion and shrinkage in the sciatic notch and femoral head region, T2D and GM fat fraction in women (N=23,364). Upper panels **a)** and **b)** show exposure PC4 of GM left and right with mediator T2D. Lower panels **c)** and **d)** show exposure T2D with mediator PC4 of GM left and right. **e)** The mean shape and the shape at the ±3 SD are displayed for PC4, showing the S2S distance variation in mm. The GM shape variations are shown in the anterior and posterior views of each GM. Direct effects are indicated by solid black arrows, whereas mediation effects are indicated by dashed grey arrows. Labels indicate effect size and p-values. Abbreviations: PC: principal component; GM: gluteus maximus; S2S: surface-to-surface; T2D: type-2 diabetes.

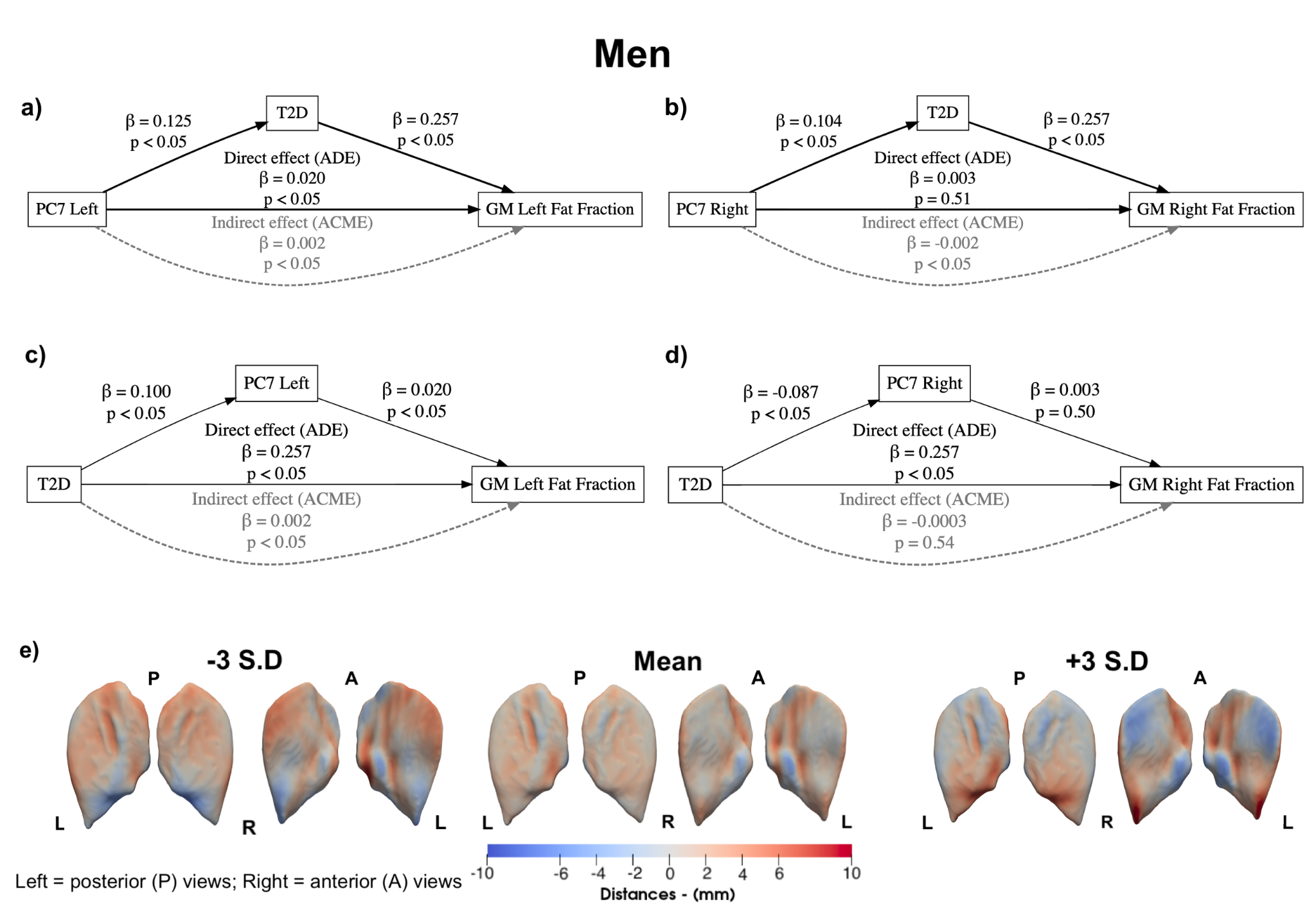

**Figure S18.** Causal mediation analysis on the relationship between PC7, explaining the left GM central/superior and femoral region shrinkage and expansion in the inferior region, T2D and GM fat fraction in women (N=23,364). Upper panels **a)** and **b)** show exposure PC7 of GM left and right with mediator T2D. Lower panels **c)** and **d)** show exposure T2D with mediator PC7 of GM left and right. **e)** The mean shape and the shape at the ±3 SD are displayed for PC7, showing the S2S distance variation in mm. The GM shape variations are shown in the anterior and posterior views of each GM. Direct effects are indicated by solid black arrows, whereas mediation effects are indicated by dashed grey arrows. Labels indicate effect size and p-values. Abbreviations: PC: principal component; GM: gluteus maximus; S2S: surface-to-surface; T2D: type-2 diabetes.

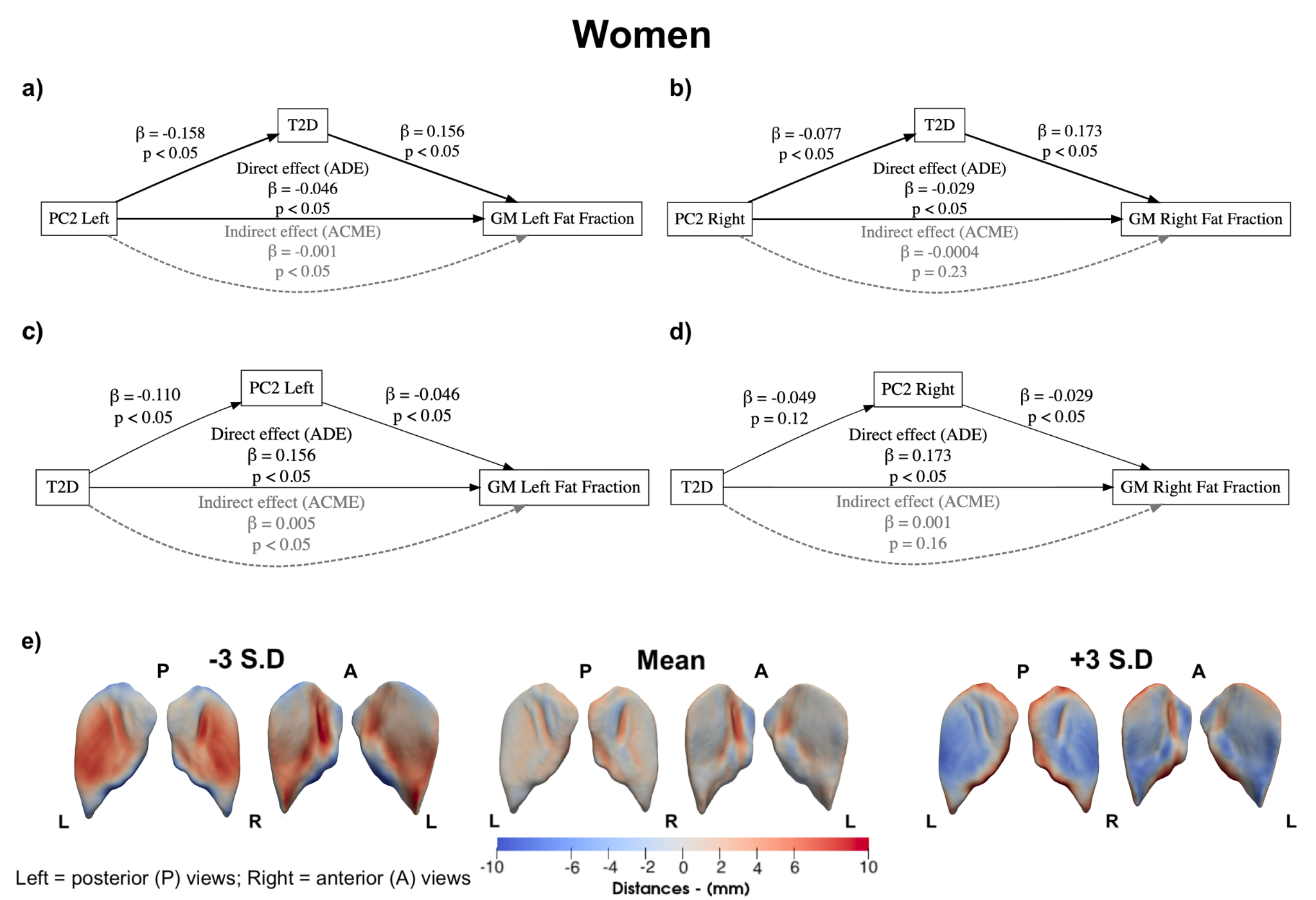

**Figure S19.** Causal mediation analysis on the relationship between PC2, explaining the left GM superior/inferior and ilioscral region expansion and central and femoral head region shrinkage, T2D and GM fat fraction in women (N=24,670). Upper panels **a)** and **b)** show exposure PC2 of GM left and right with mediator T2D. Lower panels **c)** and **d)** show exposure T2D with mediator PC2 of GM left and right. **e)** The mean shape and the shape at the ±3 SD are displayed for PC2, showing the S2S distance variation in mm. The GM shape variations are shown in the anterior and posterior views of each GM. Direct effects are indicated by solid black arrows, whereas mediation effects are indicated by dashed grey arrows. Labels indicate effect size and p-values. Abbreviations: PC: principal component; GM: gluteus maximus; S2S: surface-to-surface; T2D: type-2 diabetes.

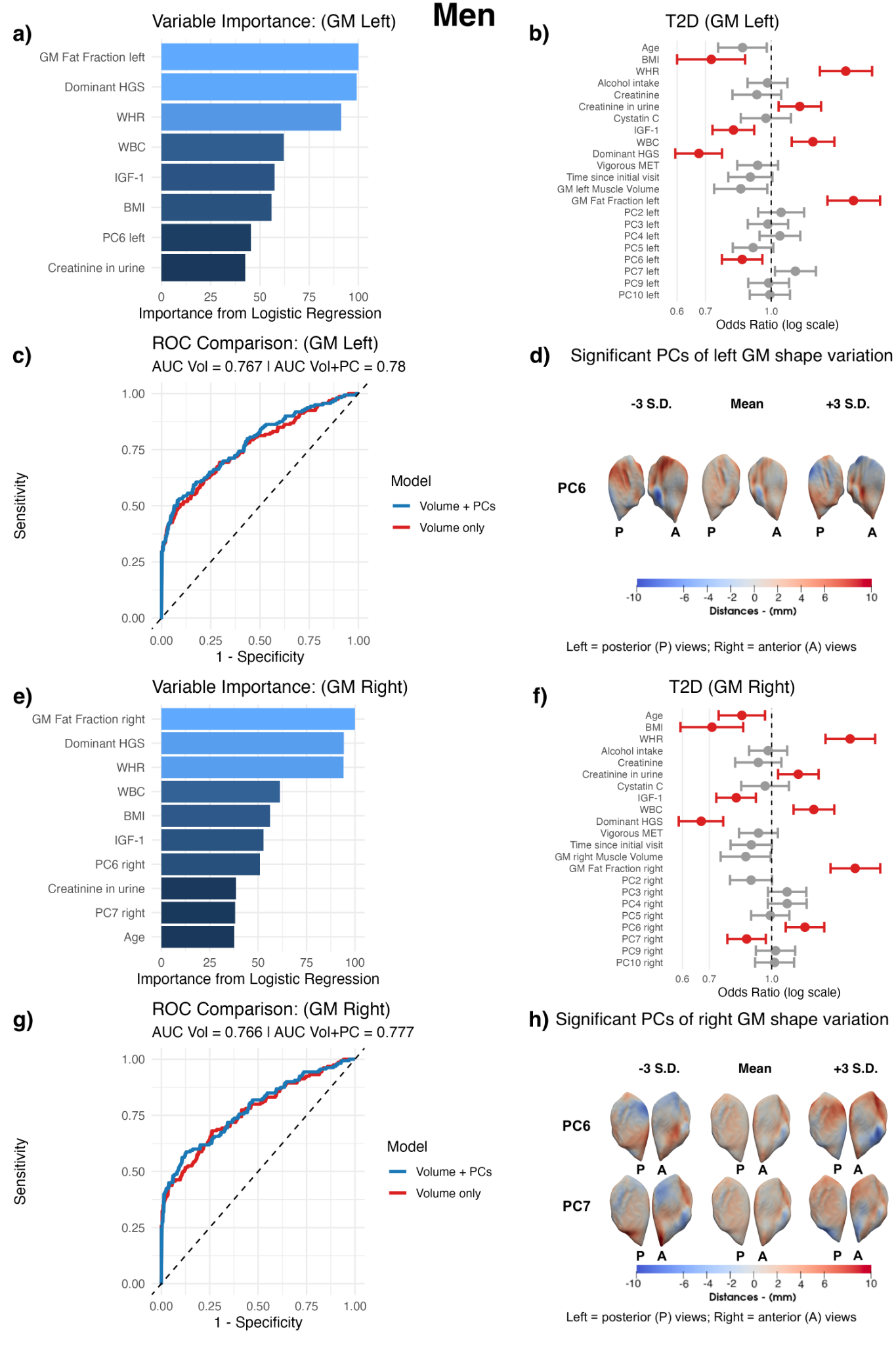

**Figure S20.** Diagnostic performance of the logistic regression model in the men case-control cohort with T2D (N=2,058) for the left GM. **a, e)** The variable importance for T2D of the volume + PCs model of the left (a) and right (e) GM. **b, f)** The odds ratios from the logistic regression for T2D for left (b) and right (f) GM adjusted for all variables in the volume + PCs model. Significant associations with FDR-adjusted p-values < 0.05 are shown in red, and non-significant associations are shown in grey. **c, g)** ROC curve comparing the volume and the volume + PCs logistic model performance for the left (c) and right (g) GM. **d, h)** The significant PCs of shape variation for the left (d) and right (h) GM. The mean GM shape and the shapes at ±3 SD are displayed for each PC, showing the S2S distance variation in mm. The GM shape variations are shown in the anterior and posterior views of the left GM. Abbreviations: BMI: body mass index; WHR: waist-to-hip ratio, IGF-1: insulin-like growth factor 1; WBC: white blood cell; HGS: hand grip strength; T2D: type-2 diabetes; GM: gluteus maximus; S2S: surface-to-surface; PC: principal component; FDR: false discovery rate; ROC: receiver operating characteristic curves.

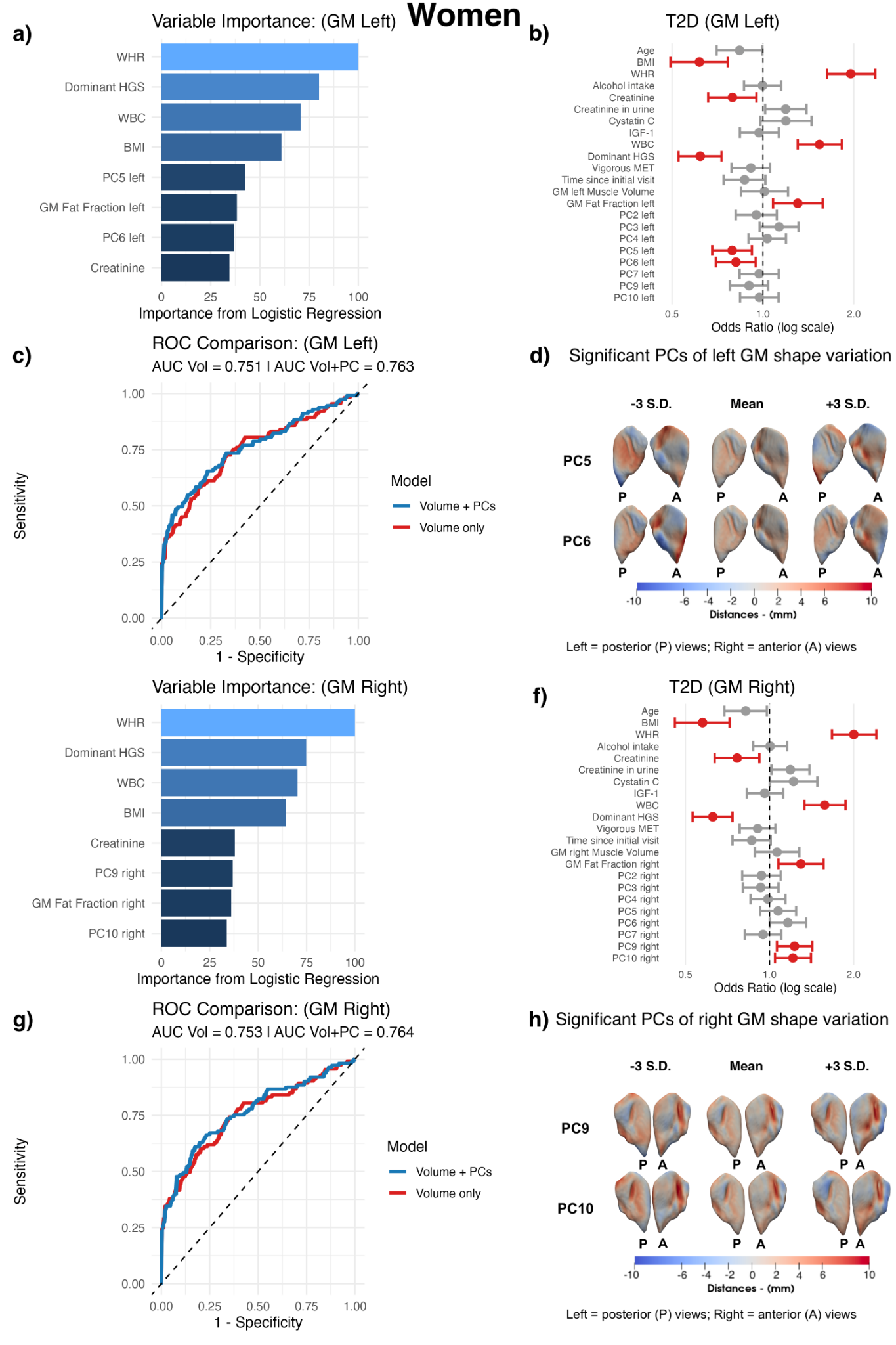

**Figure S21.** Diagnostic performance of the logistic regression model in women for the case-control cohort with T2D (N=1.318). **a, e)** The variable importance for T2D of the volume + PCs model of the left (a) and right (e) GM. **b, f)** The odds ratios from the logistic regression for T2D for left (b) and right (f) GM adjusted for all variables in the volume + PCs model. Significant associations with FDR-adjusted p-values < 0.05 are shown in red, and non-significant associations are shown in grey. **c, g)** ROC curve comparing the volume and the volume + PCs logistic model performance for the left (c) and right (g) GM. **d, h)** The significant PCs of shape variation for the left (d) and right (h) GM. The mean GM shape and the shapes at ±3 SD are displayed for each PC, showing the S2S distance variation in mm. The GM shape variations are shown in the anterior and posterior views of each GM. Abbreviations: BMI: body mass index; WHR: waist-to-hip ratio, IGF-1: insulin-like growth factor 1; WBC: white blood cell; HGS: hand grip strength; T2D: type-2 diabetes; GM: gluteus maximus; S2S: surface-to-surface; PC: principal component; FDR: false discovery rate; ROC: receiver operating characteristic curves.

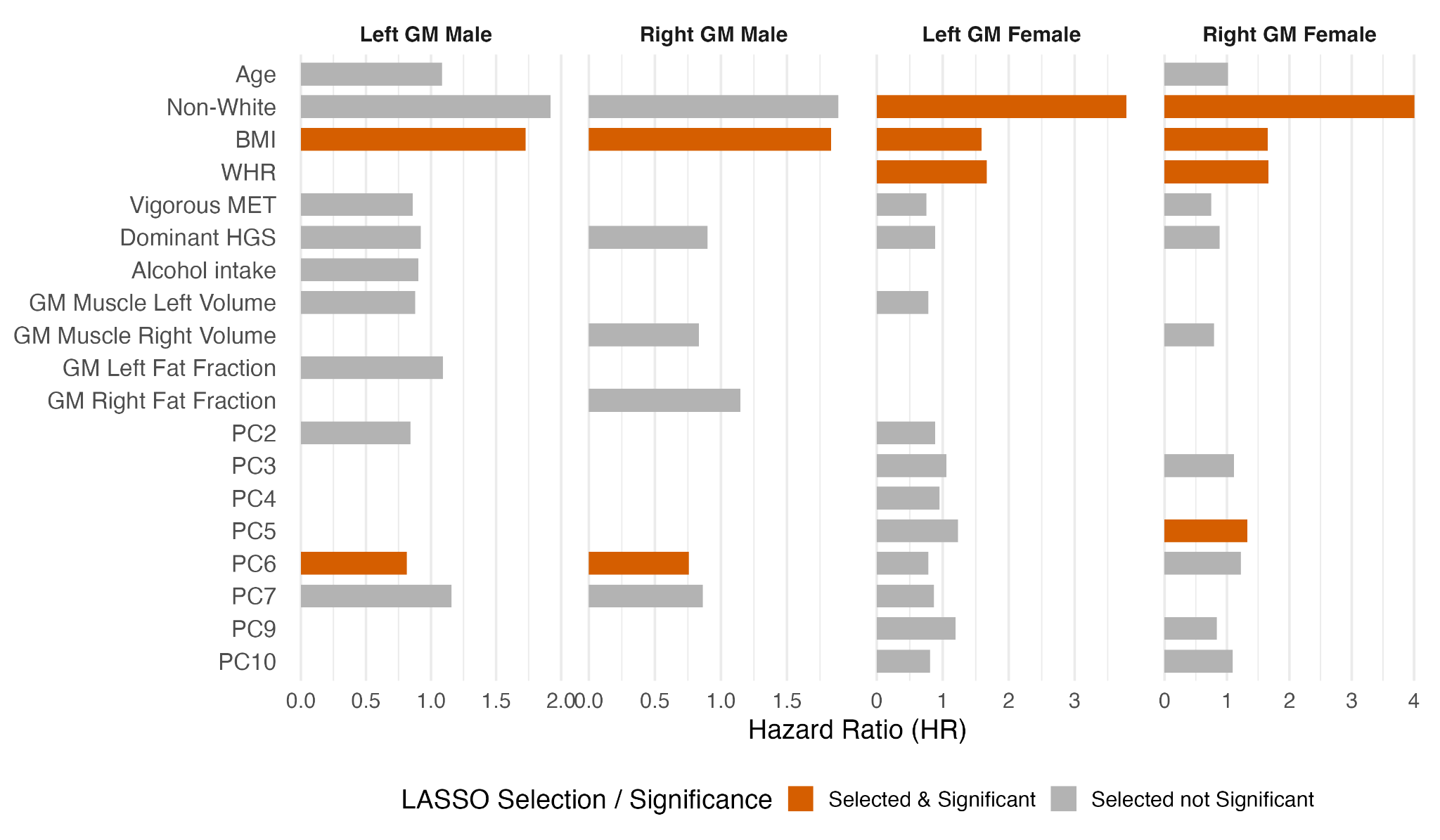

**Figure S22.** Plot showing the hazard ratios of each of the left, right GM and for all men and women participants, with all stability-selected covariates measured at the imaging visit, further using LASSO regression and 10-fold cross-validation for the T2D outcome. Red bars indicate variables selected after LASSO selection. Abbreviations: BMI: body mass index; WHR: waist-to-hip ratio, MET: metabolic equivalent task; T2D: type-2 diabetes; LASSO: least absolute shrinkage and selection operator; GM: gluteus maximus; PC: principal component.

Supplementary Videos

**Men	Women**

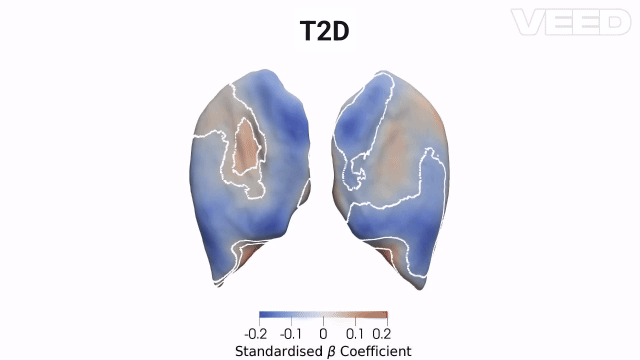

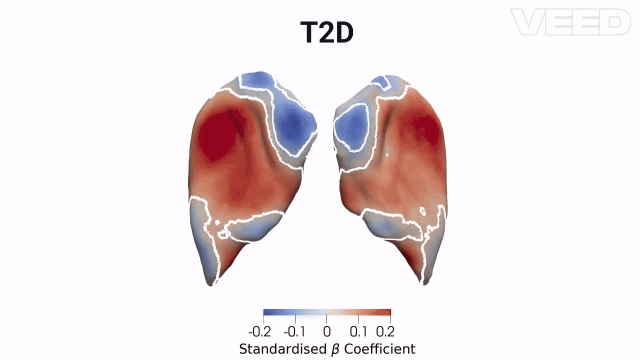

a

b

**Video S1.** Three-dimensional sex-specific SPMs of GM morphology, showing the rendering from posterior and anterior views for both left and right GM. The SPMs show the local strength of association for each disease (T2D, frailty, osteoporosis and CVD) in the model with S2S distances in **a)** men (N=23,364) and b) women (N=24,670). White contour lines indicate the boundary between statistically significant regions (p < 0.05) after correction for multiple testing, with bright red showing an outward deformation (fatty hypertrophy) and bright blue showing inward deformation (atrophy). The standardised regression coefficients () are shown with units in standard deviations for each covariate. Abbreviations: SPM: statistical parametric map; GM: gluteus maximus; S2S: surface-to-surface; T2D: type-2 diabetes; CVD: cardiovascular disease.

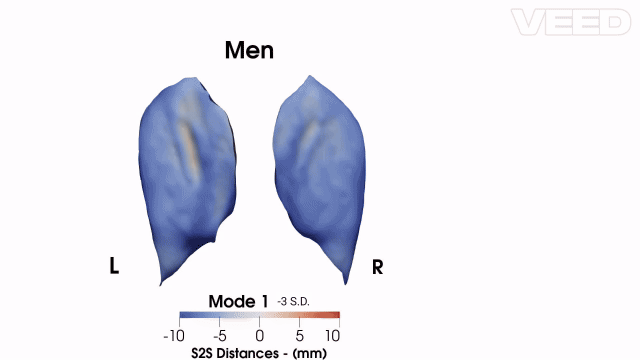

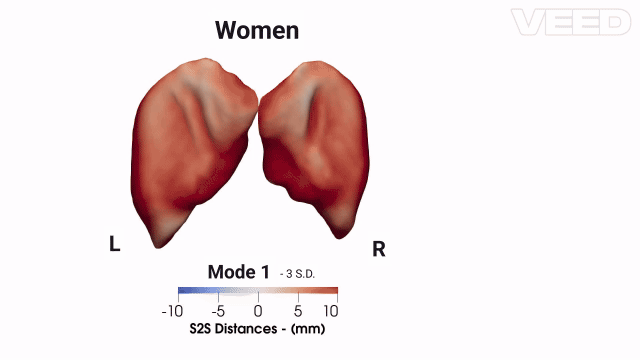

b

a

**Video S2.** The first 10 modes of shape variation for the GM S2S distances, showing posterior views for both left and right GMs in **a)** men (N=23,364) and **b)** women (N=24,670). The mean shape and the shape at the +/- 3 standard deviations are displayed for each mode, showing the GM S2S distances change in mm. For visualisation purposes, SPCA was applied jointly to two complementary representations of shape: the 3D coordinates, capturing physical shape variation, and the S2S distances, capturing computational shape differences relative to a reference surface. Together, these two components allow the modes to reflect both the physical deformation of the structure and the surface-level morphological variation. . Abbreviations: SPM: statistical parametric map; GM: gluteus maximus; S2S: surface-to-surface; SPCA: sparse principal component analysis.
